# Patterns of antibiotic cross-resistance by bacterial sample source: a retrospective cohort study

**DOI:** 10.1101/2022.03.31.22273223

**Authors:** Stacey S. Cherny, Michal Chowers, Uri Obolski

**Affiliations:** School of Public Health, Tel Aviv University, Tel Aviv, Israel; Porter School of the Environment and Earth Sciences, Tel Aviv University, Tel Aviv, Israel; Meir Medical Center, Kfar Saba, Israel; Sackler School of Medicine, Tel Aviv University, Tel Aviv, Israel

## Abstract

**Background and objectives:** Antimicrobial resistance is a major healthcare burden, aggravated when it extends to multiple drugs. While cross-resistance is well-studied experimentally, it is not the case in clinical settings, and especially not while considering confounding variables. In addition, bacteria from different sample sources may have undergone different evolutionary trajectories, therefore examining cross-resistance across sources is desirable.

**Methodology:** We employed additive Bayesian network (ABN) modelling to examine antibiotic cross-resistance in five major bacterial species, obtained from different sources (urine, wound, blood, and sputum) in a clinical setting, collected in a large hospital in Israel over a 4-year period. ABN modelling allowed for examination of the relationship between resistance to different drugs while controlling for major confounding variables.

**Results:** Patterns of cross-resistance differed across sample sources. All identified links between resistance to different antibiotics were positive, and most were present in several culture sources. However, in 15 of 18 instances, the magnitudes of the links were significantly different between sources compared. For example, *E coli* exhibited adjusted odds ratios of gentamicin-ofloxacin cross-resistance ranging from 3.0 (95%CI [2.3,4.0]) in urine samples to 11.0 (95%CI [5.2,26.1]) in blood samples.

**Conclusions and implications:** Our results highlight the importance of considering sample sources when assessing likelihood of antibiotic cross-resistance and determining antibiotic treatment regimens and policies.

**Abstract Importance:** We examine the patterns of antibiotic resistance of a given bacterial species, obtained from different clinical infection locations, while accounting for potentially relevant clinical variables. We find that such patterns of cross-resistance between pairs of antibiotics vary between culture sources (e.g., urine vs blood samples), indicating different selective pressures. These findings have implications on prescription policies aiming to minimize collateral resistance.

## Background

Antimicrobial resistance is a substantial healthcare burden, causing increased use of hospital resources, changes in treatment protocols, and excess morbidity and mortality [1]. When bacteria are resistant to multiple drugs, the problem is exacerbated. As a result, studying bacterial resistance is a major area of inquiry [2]. Many predictors of resistance in clinical settings have been identified, including age, patients’ independence status, and previous antibiotic usage [3,4]. Additionally, the interrelationship of resistance between different antibiotics is of primary clinical relevance.

Cross-resistance is the phenomenon where a bacterial isolate that is susceptible (or resistant) to a particular drug, will often be susceptible (or resistant) to a different drug. Cross-resistance of bacteria to different drugs is a commonly observed phenomenon in experimental [5–12] and clinical settings [13–17]. It is especially prevalent between antibiotics from the same antibiotic class, although differences in the drugs may lead to imperfect associations [12]. On the other hand, discordant resistance is the phenomenon where susceptibility to one drug is associated with resistance to another (and vice versa). Discordordent resistance is often demonstrated in experimental studies [5–10]. The experimental paradigm typically involves exposure of bacterial cultures to increased doses of a single antibiotic over many generations, and then testing the adapted population for resistance to other antibiotics. However, such experimental conditions do not necessarily parallel bacteria living in human populations. As a result, it is perhaps not surprising that discordant resistance is rarely observed in clinical settings [13,14]. Nonetheless, a recent paper proposed a method of analysis of clinical MIC data that is somewhat analogous to the experimental approach, that not only examines the existence of cross or discordant resistance, but attempts to establish direction of effect [14]. They found some of the limited discordant (and cross) resistance present was unidirectional.

Revealing cross-resistance patterns in clinical settings is not trivial. Observational data are often subject to confounding due to non-random exposure antibiotics, which can substantially mask the underlying cross-resistance patterns. We have previously employed additive Bayesian network (ABN) modelling to deal with such confounding in clinical data [15]. ABN models allow uncovering the association structure among a set of variables, while controlling for relevant confounders. However, the differences of cross-resistance patterns between sample sources have not been explored in depth previously, while adjusting for confounders. Bacteria from different sample sources may have undergone divergent evolutionary trajectories as a result of different selective forces, perhaps leading to distinct patterns of resistance. Such differences can have clinical implications on short- and long-term antibiotic treatment selection.

In this study, we use a large clinical dataset obtained from an Israeli hospital over a 4-year period, to study cross-resistance in different sample sources. We employ ABN modelling to control for potential confounding variables and explore the resistance network structures by bacterial species and sources of the bacterial culture. This allows for unbiased examination of whether relationships between drug resistance differs by culture source.

## Methodology

### Data

We obtained data pertaining to all positive bacterial cultures drawn in Meir Medical Center, Israel, a 740 bed hospital with approximately 60,000 admissions per year, from 2016-01-02 to 2019-12-31. The corresponding medical history, demographics, previous hospitalizations, and previous in-hospital antibiotic usage in the year prior to the infection, of patients from whom the cultures were drawn, were also available. Bacterial cultures were tested for antibiotic resistance for an array of antibiotics, and results of non-susceptibility and resistance were combined into a ‘resistant’ category. All antibiotics tested were systemic, with the exception of mupirocin, which is topical. Nonetheless, given we had data on bacterial resistance to mupirocin, it was still included in the analyses. Bacterial infections were considered nosocomial if cultures were drawn >48 hours after admission. A summary of these variables is presented in Tables 1 and 2. For our analyses, we selected the five bacterial species with the largest sample sizes available in the dataset: *Escherichia coli, Staphylococcus aureus, Pseudomonas aeruginosa, Klebsiella pneumoniae*, and *Proteus mirabilis*, in order of decreasing frequency.

**Table 1:** Descriptive statistics of patients for *E coli* and *K pneumoniae* bacterial isolates, by source of sample

|  | *E coli* |  |  | *K pneumoniae* |  |  |
| --- | --- | --- | --- | --- | --- | --- |
|  | Urine | Wound | Aerobic blood | Urine | Wound | Aerobic blood |
|  | (N=2546) | (N=596) | (N=383) | (N=798) | (N=219) | (N=108) |
| Age [mean (SD)] | 70.4<br>(21.8) | 62.8<br>(19.7) | 74.6<br>(15.4) | 74.4<br>(17.2) | 64.4<br>(17.3) | 71.6<br>(15.4) |
| Female (%) | 1768<br>(69.4) | 252<br>(42.3) | 232<br>(60.6) | 465<br>(58.3) | 86 (39.3) | 39 (36.1) |
| Immunosuppression (%) | 89 (3.5) | 23 (3.9) | 24 (6.3) | 43 (5.4) | 9 (4.1) | 6 (5.6) |
| Dementia (%) | 199 (7.8) | 10 (1.7) | 23 (6.0) | 65 (8.1) | 1 (0.5) | 8 (7.4) |
| Diabetes (%) | 386<br>(15.2) | 97 (16.3) | 61 (15.9) | 157<br>(19.7) | 57 (26.0) | 26 (24.1) |
| COPD (%) | 110 (4.3) | 14 (2.3) | 26 (6.8) | 32 (4.0) | 9 (4.1) | 10 (9.3) |
| CRF (%) | 138 (5.4) | 19 (3.2) | 23 (6.0) | 52 (6.5) | 20 (9.1) | 7 (6.5) |
| BMI >30 (%) | 119 (4.7) | 18 (3.0) | 22 (5.7) | 38 (4.8) | 6 (2.7) | 4 (3.7) |
| Cephalosporin use (%) | 855<br>(33.6) | 314<br>(52.7) | 152<br>(39.7) | 369<br>(46.2) | 140<br>(63.9) | 59 (54.6) |
| Non-cephalosporin beta-lactam use (%) | 253 (9.9) | 192 (32.2) | 46 (12.0) | 129 (16.2) | 80 (36.5) | 24 (22.2) |
| Beta-lactam & betalactamase inhibitor use (%) | 209 (8.2) | 118 (19.8) | 34 (8.9) | 98 (12.3) | 57 (26.0) | 17 (15.7) |
| Nitroimidazole use (%) | 167 (6.6) | 197 (33.1) | 56 (14.6) | 92 (11.5) | 72 (32.9) | 18 (16.7) |
| Aminoglycoside use (%) | 131 (5.1) | 70 (11.7) | 17 (4.4) | 69 (8.6) | 22 (10.0) | 6 (5.6) |
| Fluoroquinolone use (%) | 84 (3.3) | 40 (6.7) | 7 (1.8) | 40 (5.0) | 14 (6.4) | 6 (5.6) |
| Other drug family use (%) | 177 (7.0) | 56 (9.4) | 22 (5.7) | 79 (9.9) | 29 (13.2) | 16 (14.8) |
| Days hospitalized [Median (IQR)] | 0.0125 (4.07) | 0.083 (5.86) | 0.126 (5.64) | 0.185 (7.06) | 1.07 (7.50) | 0.321 (8.87) |
| Nosocomial (%) | 548 (21.5) | 366 (61.4) | 84 (21.9) | 246 (30.8) | 146 (66.7) | 45 (41.7) |
| Polymicrobial (%) | 142 (5.6) | 425 (71.3) | 32 (8.4) | 130 (16.3) | 173 (79.0) | 14 (13.0) |
| CAZ resistance (%) | 860 (33.8) | 168 (28.2) | 93 (24.3) |  |  | 31 (28.7) |
| GEN resistance (%) | 375 (14.7) | 102 (17.1) | 37 (9.7) | 166 (20.8) | 51 (23.3) | 21 (19.4) |
| OFX resistance (%) | 869 (34.1) | 148 (24.8) | 95 (24.8) | 97 (12.2) | 28 (12.8) | 12 (11.1) |
| SXT resistance (%) | 882 (34.6) |  |  | 331 (41.5) |  |  |
| TZP resistance (%) |  | 22 (3.7) |  | 31 (3.9) | 12 (5.5) |  |

Note: shaded values represent variables omitted from the model for practical reasons.
COPD, chronic obstructive pulmonary disorder; CRF, chronic renal failure; CAZ, ceftazidime; GEN, gentamicin; OFX, ofloxacin; SXT, sulfamethoxazole/trimethoprim, TZP, piperacillin/tazobactam.

**Table 2:**
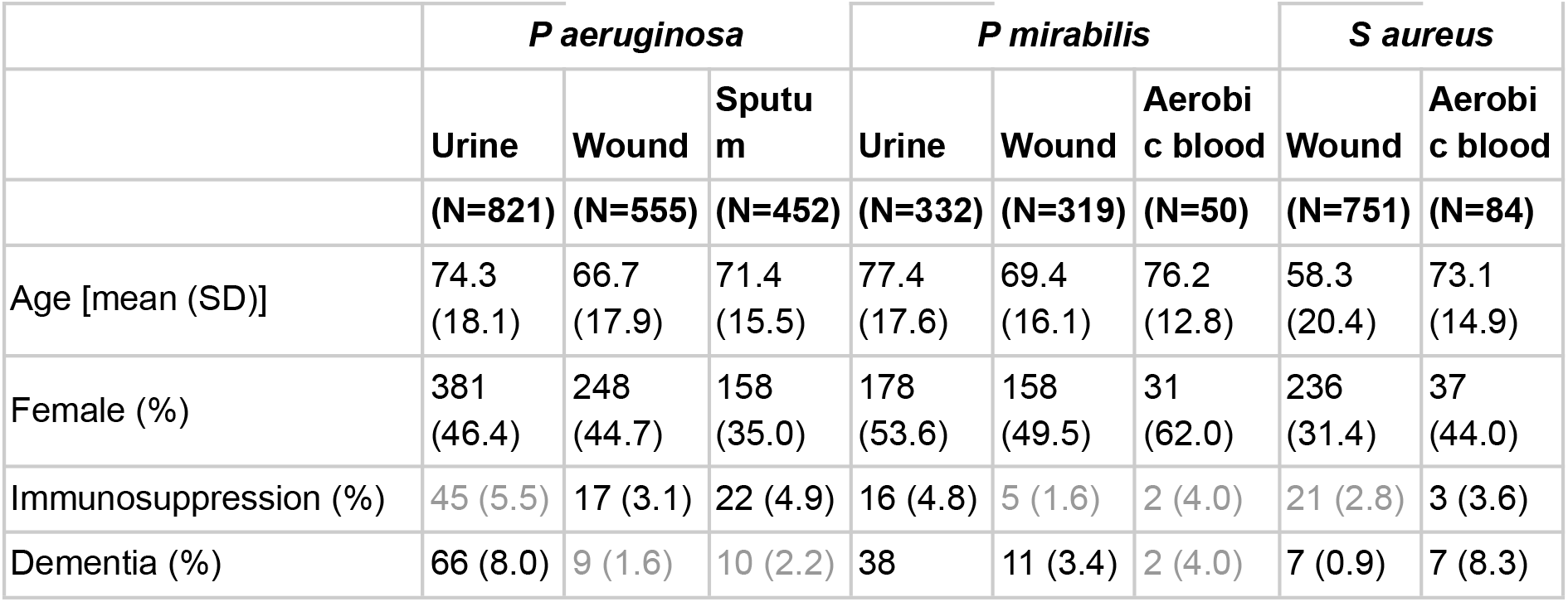
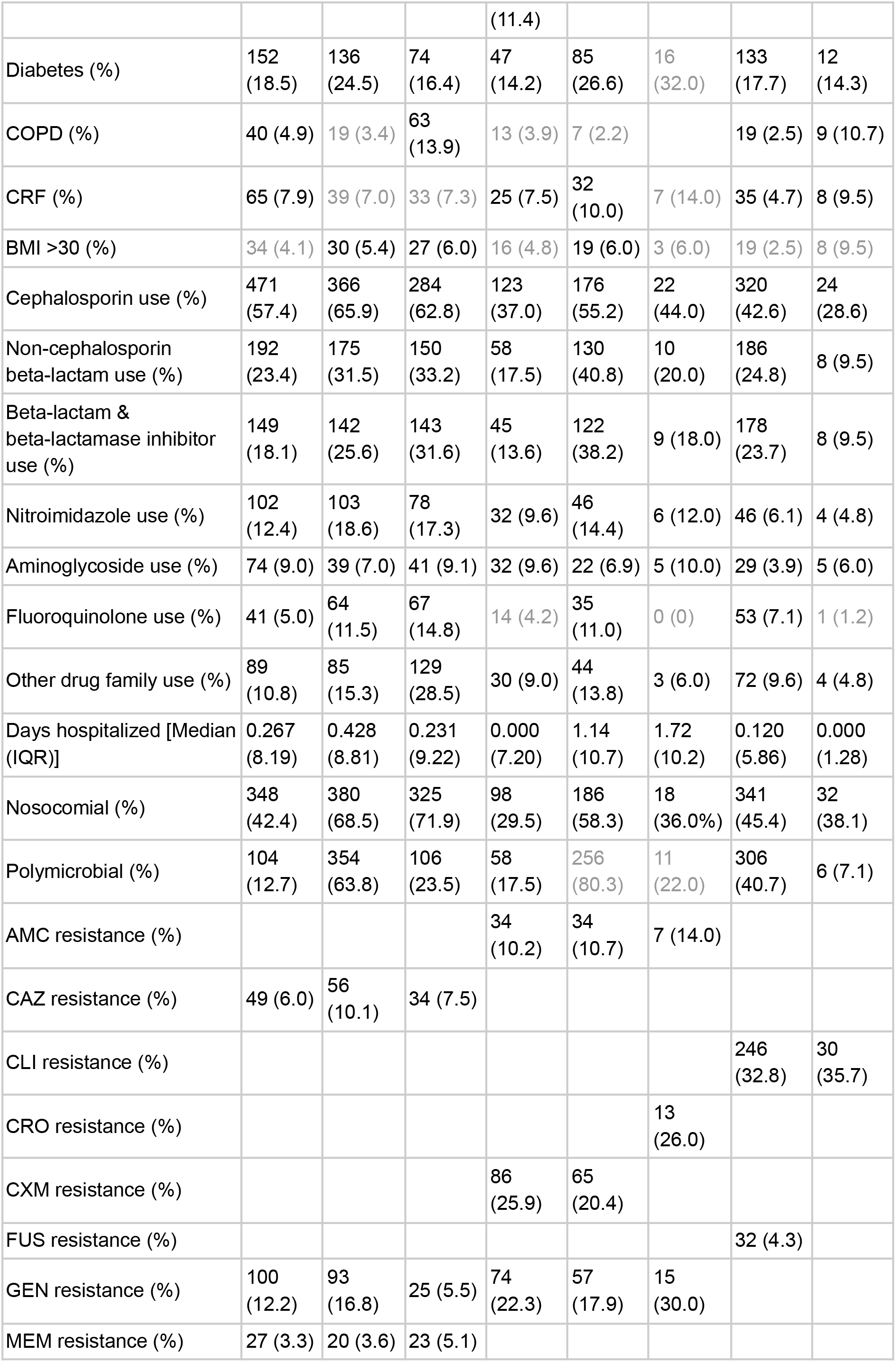
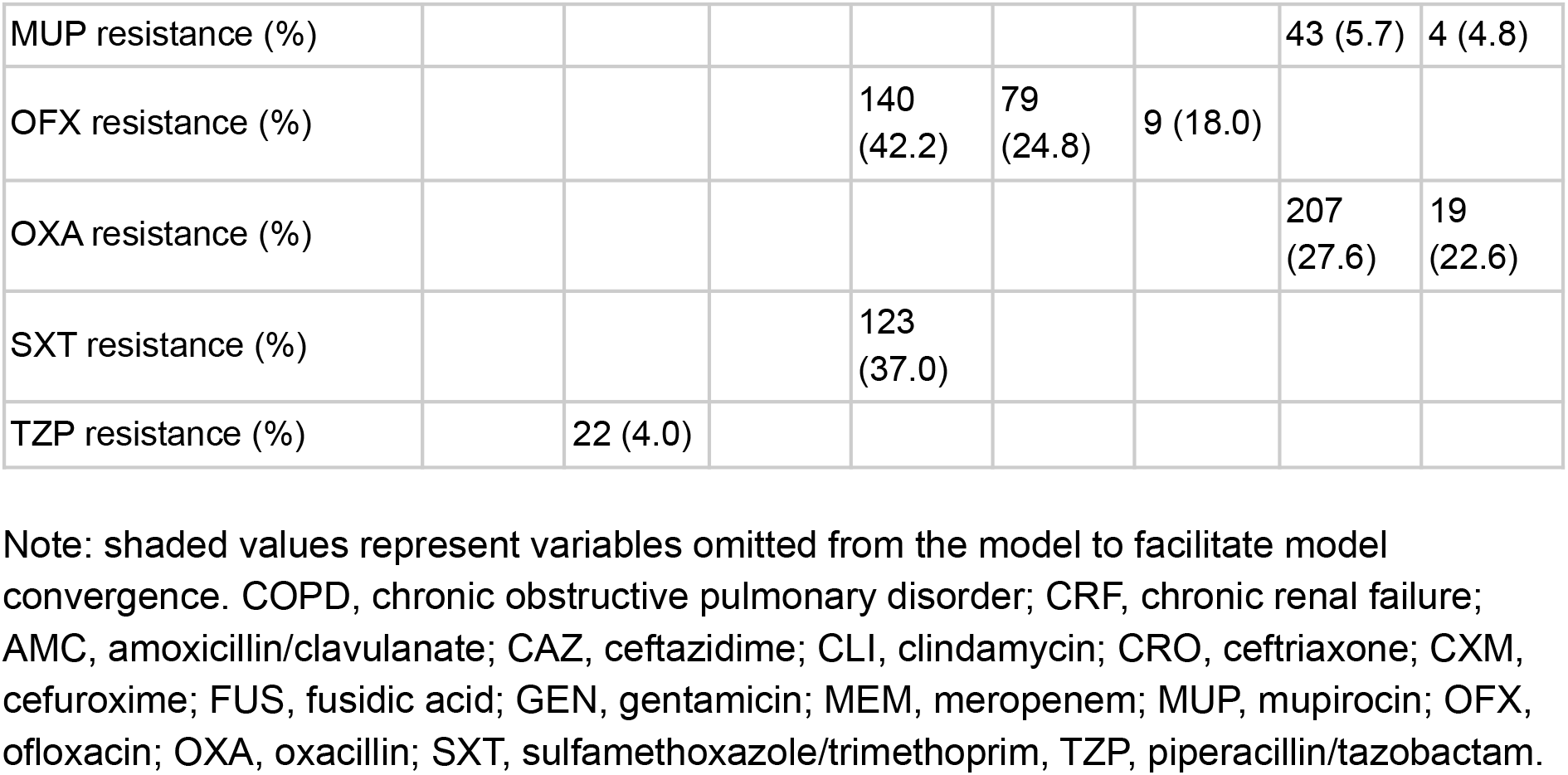
Descriptive statistics of patients for *P aeruginosa, P mirabilis*, and *S aureus* bacterial isolates, by source of sample

### Statistical analysis

We selected which antibiotics to include in the analysis by keeping only those with minimal missing data and which did not reduce the number of complete cases appreciably (<10% loss). We performed some variable selection to assure stable statistical models with no perfect or near-perfect separation, by not including perfectly or near perfectly correlated antibiotics and selecting only antibiotics which contained a minimum of 3% resistance in each bacterial subsample. Antibiotics excluded from analysis due to high collinearity with included variables are presented in Supplementary Table S1, along with their range of tetrachoric correlations with the relevant included variables, across the various sample sources. Excluded antibiotics were usually, but not always, of the same drug family. For example, while ceftazidime, ceftriaxone, cefuroxime, and cefalexin are all cephalosporins and we often removed three of these four, ampicillin, a beta-lactam, was removed due to high correlation with the cephalosporins. This resulted in analysis of between three and five antibiotics for the five bacterial species cultured from the various sources, each analyzed separately.

When constructing the ABN, the following covariates were included, in addition to the antibiotic resistance tests: demographic variables (*age, sex*, and *days hospitalized* in the previous year), presence of five medical conditions (*immunosuppression, dementia, diabetes*, chronic obstructive pulmonary disorder (*COPD*), chronic renal failure (*CRF*), and *obesity* (BMI over 30), binary culture type variables (*nosocomial* and *polymicrobial*), and a binary variable for antibiotic use in hospital in the previous year.

Antibiotics used were first grouped into drug families and the six most frequent families across the entire dataset were included, along with whether any other antibiotics were taken (which did not belong to the six largest families). The six families were (in descending frequency of use) cephalosporins, beta-lactams (that are not cephalosporins), beta-lactamase inhibitors, nitroimidazoles, aminoglycosides, and fluoroquinolones, plus a seventh category including all other antibiotics.

Table 1 presents summary statistics for *E coli* and *K pneumoniae*, for all variables used in our models, and Table 2 presents summary statistics for *P aeruginosa, P mirabilis, and S aureus*. For some models, certain covariates needed to be excluded from the analysis in order for the ABN models to converge due to insufficient sample size. In addition, for the *E coli* urine analysis, to allow the model to run within a reasonable amount of time (days rather than months), two variables were omitted which did not have a direct connection with resistance in the initial search. Excluded variables are noted in Tables 1 and 2.

Data were analyzed using ABN modelling,[18,19] with version 2.5.0 of the R package abn[20] on an R 4.1.0 installation.[21] Briefly, ABN modelling is a purely data-driven, exploratory approach, originating in machine learning, that is often used for hypothesis generation for causation among a set of variables. By searching likely networks linking a set of variables, a model for the dependency of the variables in the data can be inferred without making strong prior assumptions. This model shows which variables are directly connected, or linked, via arcs, and the coefficients of these arcs are directly analogous to the adjusted odds ratios obtained from multiple logistic regression analysis.[18]

While no assumptions of causal relationships are required, we restricted the model space to disallow causal paths that made no sense for our study, details of which are included in the Supplementary Methods. We followed a multi-step procedure: first identifying the most-likely network structure, using the abn package; then performing a parametric bootstrap, using JAGS to simulate 1000 datasets per model; then using ABN to reanalyze each simulated dataset, to correct for overfitting; and finally, re-estimating parameters using ABN, only allowing for connections seen in at least 50% of the analyses of the simulated datasets, and computing 95% Bayesian credible intervals (CIs) for each of the parameter estimates. The procedure we employed has been previously described and applied to antibiotic resistance data by us.[15] One difference in the method of analysis in the present paper is that in order to speed up the parametric bootstrap, we dropped all variables that had no connection to any other variable in the initial network and reran the full procedure on the reduced dataset. This would have no effect on the results presented here. In addition, we limited the search for the maximum number of parents to eight, due to prohibitive computational time needed to conduct the search allowing more parents. Supplementary Table S2 contains the number of arcs present in the model both before and after bootstrapping, with the one model limited to 8 without reaching a plateau noted.

To test whether arcs present in different sources of the same bacterial species were significantly different in magnitude, we randomly sampled from the posterior distributions of a given arc and computed the difference between a random pair posterior ln(OR) estimates, which yielded a distribution of ln(OR) differences. From that, we present the 95% CIs of the differences and examine whether zero was inside the intervals, indicating no significant difference in arc parameter estimates. The posterior estimates were computed at a granularity of 10,000 per parameter and 100,000 pairs were sampled with replacement to generate the empirical distribution of ln(OR) differences.

### Ethics

The study was approved by the Institutional Review Board (Helsinki) Committee of Meir Medical Center. Since this was a retrospective study, using archived medical records, an exemption from informed consent was granted by the Helsinki Committee.

## Results

In Figure 1 we present an example of an ABN estimated for *P aeruginosa* in a wound sample. The antibiotic resistance tests had direct links to *sex, polymicrobial infection*, and previous use of fluoroquinolones, beta-lactam inhibitors, and less-common antibiotics (*other)*. However, resistance to MEM was not linked to any variable. Thus, the network yields less biased associations between the different antibiotics by controlling for various patient covariates. ABNs for all bacterial species investigated are presented in Supplementary Figures S1-S14, with parameter estimates and 95% CIs presented in Supplementary Tables S3-S16.

**Figure 1:**
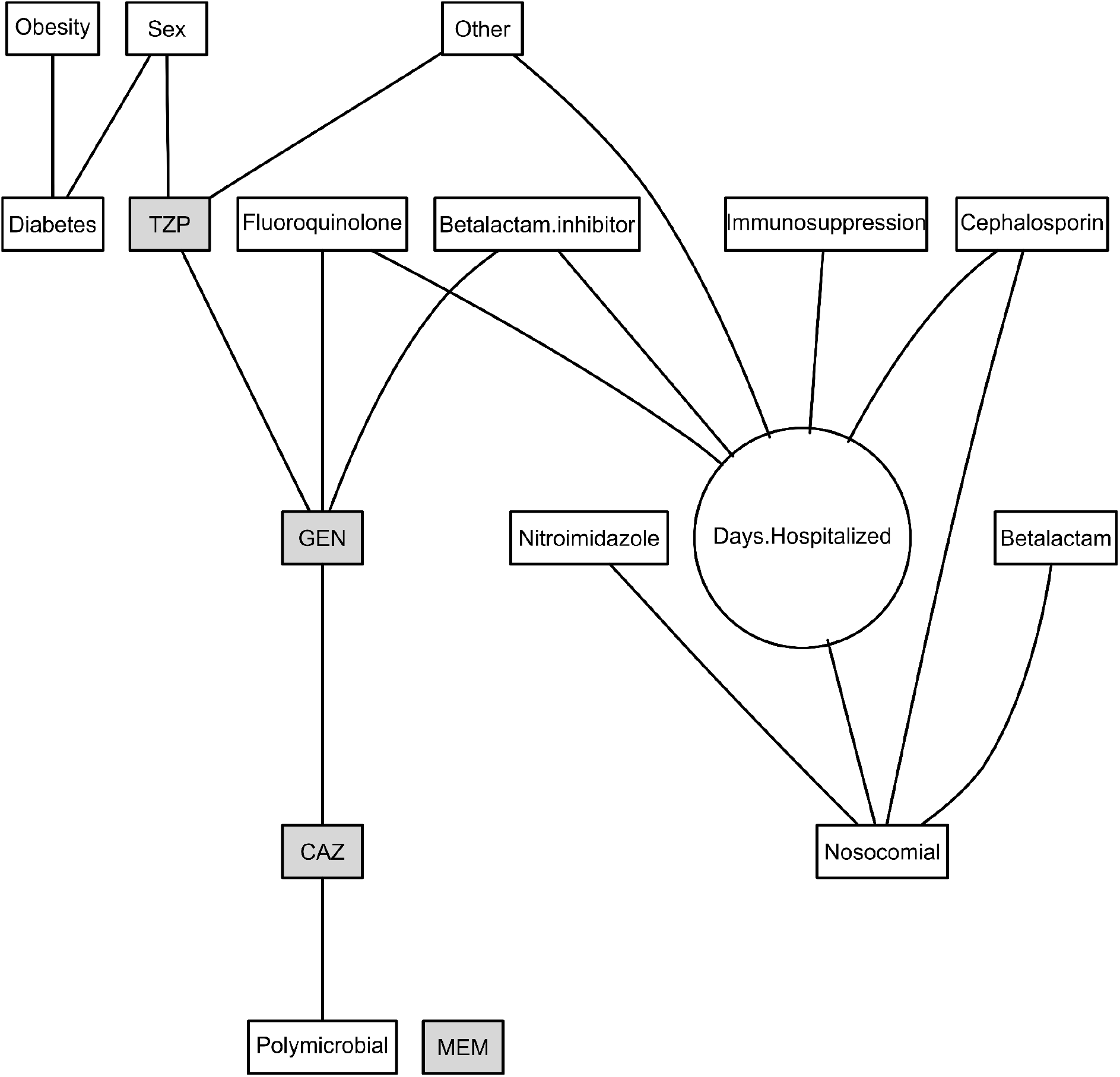
Additive Bayesian network for *P aeruginosa* in wound samples. Variables in rectangles are binary and the one in a circle is continuous. Drug family variables refer to a particular drug taken in the prior year. Antibiotic resistance tests are shown in shaded boxes: CAZ, ceftazidime; GEN, gentamicin; MEM, meropenem, TZP, piperacillin/tazobactam.

To better summarize our results, we present consensus graphs (Figure 2). These graphs present the relationships among drug resistances from all bacterial sources for a given bacterial species. Figure 2 contains 5 panels, A-E, representing the relationships among the antibiotic resistance tests in *E coli, K pneumoniae, P aeruginosa, P mirabilis, and S aureus*, respectively, obtained from fitting the ABN models (as the one presented in Figure 1; see Supplementary). The nodes in the consensus graphs are denoted with both the antibiotic resistance tests included in the ABN models and corresponding antibiotics excluded due to very high correlations with the included antibiotic, as seen from examination of the pairwise tetrachoric correlations (Supplementary Table 1).

**Figure 2:**
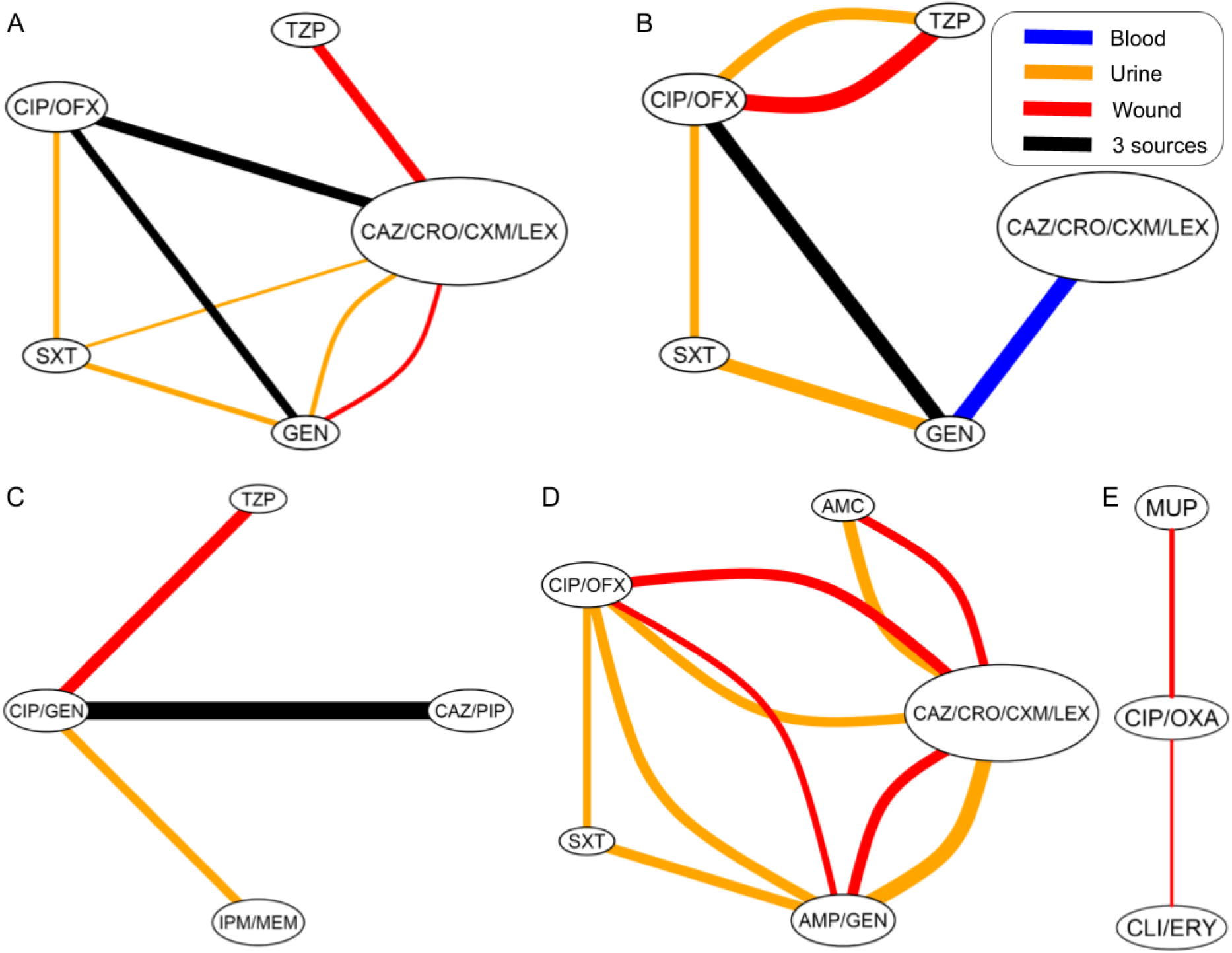
Consensus network graphs for the five bacterial species: *E coli* (A),*K pneumoniae* (B), *P aeruginosa* (C), *P mirabilis* (D), and *S aureus* (E). Line color depicts in which bacterial source model the link is present, with line width proportional to the ln(OR) (or mean ln(OR) for black lines). Orange=urine; red=wound; blue=aerobic blood; black=urine, blood, and wound, except in *P aeruginosa*, where black denotes urine, blood, and sputum. AMC, amoxicillin/clavulanate; AMP, ampicillin; CAZ, ceftazidime; CIP, ciprofloxacin; CRO, ceftriaxone; CXM, cefuroxime; ERY, erythromycin; GEN, gentamicin; IPM, imipenem; LEX, cefalexin; MEM, meropenem; MUP, mupirocin; OFX, ofloxacin; OXA, oxacillin; PIP, piperacillin; TZP, piperacillin/tazobactam; SXT, sulfamethoxazole/trimethoprim.

In Figure 2 arcs are color-coded by sample source: orange arcs are in urine, red in wound, blue in aerobic blood, and black for arcs present in all three sources for a particular bacterial species. In all four of the species where ciprofloxacin and gentamicin were tested, they were linked, with the exception of *P mirabilis*, where they were linked in two of the three sample sources. Ceftazidime and gentamicin were also linked in four species: in *P aeruginosa*, in all three sources and in two of three sources in *E coli* and *P mirabilis*. Piperacillin/tazobactam was directly linked to ciprofloxacin in two of the three species for which it was tested and in two sample sources of *K pneumoniae*. Importantly, all direct connections were consistently positive throughout all antibiotics and all bacterial species.

Finally, we sought to analyse whether the magnitude of the cross-resistance links differed between sample sources. Parameter estimates of the cross-resistance links, along with their 95% CIs, are presented in Figure 3 for those parameters estimated in more than one sample source of a particular bacterial species. The median differences between these parameter estimates in different sample sources, along with 95% CIs for the difference, within bacterial species, are presented in Supplementary Table S17. While many arcs were present in more than one culture source for a given species, only in three of 18 instances were the magnitudes of those parameter estimates not significantly different between sources compared. In *E coli*, the link between ofloxacin and ceftazidime was not significantly different in aerobic blood than in urine, as was the link between ceftazidime and gentamicin in urine versus wound. The third pair of estimates not significantly different was cefuroxime and ofloxacin in the *P mirabilis* cultures from urine versus wound. All other links present in two different culture sources were found significantly different between sources, as determined by the 95% CIs of the differences not including zero.

**Figure 3:**
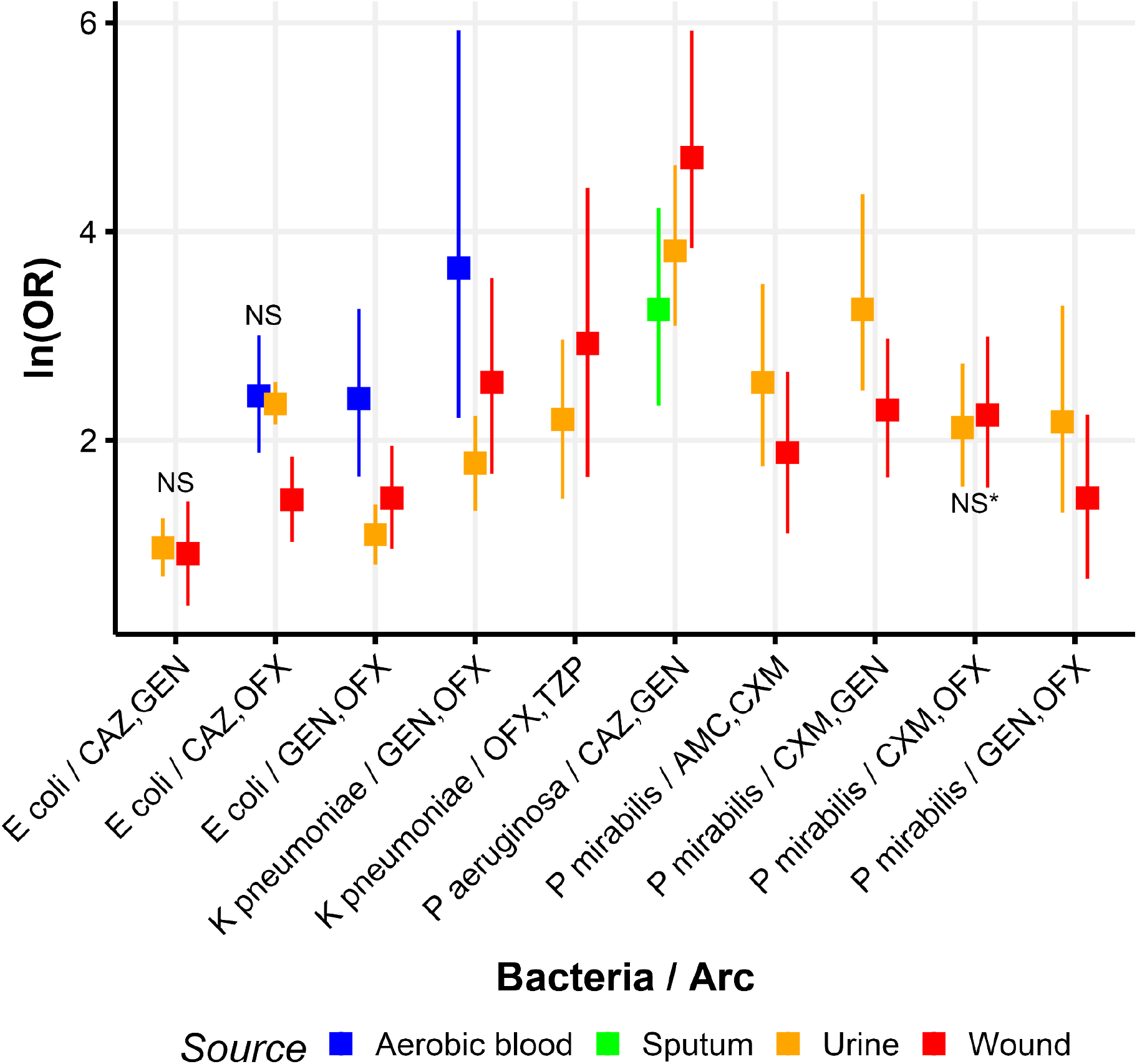
Parameter estimates (ln(OR)) and their 95% credible intervals, for arcs between pairs of antibiotics, by bacteria and sample source, where both antibiotics appear in more than one sample source for a given bacteria. NS near a pair denotes that the two estimates were not significantly different, with * denoting within .01 from the null. The 95% CI of the estimate of the difference between all other pairs tested within a bacterial species did not contain zero. Line color depicts in which bacterial source the estimate was obtained: orange=urine; blue=aerobic blood; red=wound; green=sputum. AMC, amoxicillin/clavulanate; CAZ, ceftazidime; CIP, ciprofloxacin; CXM, cefuroxime; GEN, gentamicin; OFX, ofloxacin; TZP, piperacillin/tazobactam. *The 95% credible interval contains zero to one decimal place.

## Conclusions and implications

Bacterial cross-resistance has been studied in experimental [5–11] and clinical settings [13–17]. However, to the best of our knowledge, cross-resistance patterns have not been compared between clinical sample sources, while controlling for biases arising from the retrospective nature of such data. In this work, we examined patterns of cross-resistance across bacterial culture sources, while adjusting for potential confounders, using ABNs. We found that the patterns and magnitude of cross-resistance among pairs of antibiotics may vary significantly between sample sources.

The general patterns of cross-resistance in our study are consistent with previous research. For example, all identified links were positive; and links within antibiotic classes, with similar resistance mechanisms, were high. In fact, the cross-resistance estimates of some antibiotics could not be disentangled in our analysis, due to their high correlations. Such a grouping due to high correlations was formed by the cephalosporins ceftazidime, ceftriaxone, cefuroxime, and cefalexin in *E coli, K pneumoniae, and P mirabilis*, for all tested sample sources. As expected, resistance to older generations of cephalosporins did not necessitate resistance to newer generations, but almost never vice versa. However, because those instances were relatively rare, the correlations between antibiotics remained very high. Some discrepancies were also found between our study and a large and recent study of cross-resistance in clinical bacterial samples [13]. The differences between the results could be due to several major differences between the analyses. Firstly, we controlled for an array of relevant covariates using ABNs. Secondly, we examined relationships among resistances stratified by sample sources. Finally, our data are relatively homogenous, since they originated from a single center. In contrast, the data used in the mentioned study originated from over 35 separate hospitals across the US.[13]

While all links identified in our study were positive or non-existent, 15 of 18 pairwise links were significantly different in their magnitude between different sample sources. This suggests that the extent of cross resistance found might depend on the selective pressures exerted by the environment the bacteria were sampled from. In several cases, these differences were dramatic. For instance, in *K pneumoniae*, the OR between gentamicin and ofloxacin ranged from around 6 in urine to nearly 40 in blood. In *E coli*, the same antibiotics varied in their ORs from 3 in urine to 11 in blood. Such differences can distinguish between a treatment being merely unlikely to succeed, given one type of resistance, to being almost futile.

A limitation of our analyses is that sample size differed greatly both in bacterial species and sample sources. This translates into differing levels of statistical power across ABN analyses. For example, we observe the most arcs in the network for *E coli* obtained from urine, potentially because the sample size was largest in that analysis. For the three other bacterial species cultured from urine, it is apparent that there are more arcs in the models connecting antibiotic resistance tests as compared to other culture sources, again potentially due to urine having the largest sample sizes. Nonetheless, while sample sizes were modest in some groups, ample data was available for others, and the study examined all bacterial cultures obtained in a large hospital over a four year period, making it quite comprehensive. Furthermore, the patterns found in our analyses are not necessarily generalizable to other countries or even hospitals. Resistance patterns are determined by prescription patterns, patient demographics and a host of other factors, and are expected to differ across time and space.[4,22] However, our results of different magnitudes of cross-resistance between sample sources have a biological rationale, and we therefore expect them to be more generalizable.

To conclude, using a large clinical dataset obtained from admissions to a major hospital in Israel, over a 4-year period, we applied ABN modelling to control for potential confounding variables and explored resistance network structures stratified by bacterial species and source of the bacterial culture. This allowed for less biased examination of the relationships between drug resistance and culture sources. We found similar patterns of the existence of cross-resistance in different culture sources within bacterial species; however, the magnitude of those relationships differed substantially between sample sources. This highlights the importance of considering sample sources when assessing the likelihood of antibiotic cross-resistance. Future antibiotic prescription policies aiming to minimize collateral resistance should therefore be differentially determined by the source of infection.

## Data Availability

Data are proprietary but can be made available upon reasonable request from the authors.

## Transparency declarations

None to declare.

## Data sharing

Data are proprietary but can be made available upon reasonable request from the authors.

