## Supplementary Material for "Patterns of antibiotic cross-resistance by bacterial sample source: a retrospective cohort study"

### Supplementary Methods

While no assumptions of causal relationships are required, we restricted the model space to disallow causal paths that made no sense for our study: (1) *sex* could only cause other variables and not be caused by any other variable, (2) *age* at testing could be caused by *sex* but no other variable, (3) the seven variables for antibiotics given in the previous year could only be caused by *age* and *sex* and no other variable, (4) *days hospitalized* the prior year could not be caused by presence of nosocomial infection but could be caused by any other variable, (5) *nosocomial* infection could not be caused by resistance to any drug but can be caused by any other variable; (6) *polymicrobial* cultures as well as all drug antibiotic resistance tests could be caused by any of the variables; (7) the six medical conditions can be caused by age and sex; (7) *immunosuppression* and *CRF* can be caused by any of the other medical conditions; (8) high *BMI* can only be caused by one of the other medical conditions, *dementia*; (9) *dementia*, *diabetes*, and *COPD* can be caused by each other or high *BMI* but not *immunosuppression* and *CFR*; and (10) high *BMI* can only be caused by *dementia* but not the other 4 medical conditions. In addition, no causal paths were forced to be in the model.

### Supplementary Results

**Table S1: Antibiotic resistance tests excluded from analysis due to redundancies or missing data, along with their tetrachoric correlations with highly-correlated included variables**

| Dataset | Antibiotic removed | Antibiotics it is correlated with | Tetrachoric correlation range |
|---|---|---|---|
| *E.coli* | AMP | CAZ, CRO, CXM, LEX | 0.80 - 0.93 |
| *E.coli* | CRO | CAZ | 1.00 |
| *E.coli* | CXM | CAZ | 0.99 - 1.00 |
| *E.coli* | LEX | CAZ | 0.98 - 0.99 |
| *K. pneumoniae* | AMC | CAZ, CIP, CRO, CXM, GEN, LEX, TZP | 0.70 - 0.99 |
| *K. pneumoniae* | CIP | OFX | 0.89 - 1.00 |
| *K. pneumoniae* | CRO | CAZ | 0.98 - 1.00 |
| *K. pneumoniae* | CXM | CAZ | 0.99 - 1.00 |
| *K. pneumoniae* | LEX | CAZ | 0.98 - 0.99 |
| *P. aeruginosa* | CIP | GEN | 0.88 - 0.98 |
| *P. aeruginosa* | PIP | CAZ, TZP | 0.90 - 0.99 |
| *P. aeruginosa* | IPM | MEM | 0.94 - 0.99 |
| *P. mirabilis* | AMP | CRO, CXM, GEN | 0.87 - 1.00 |

| | | | |
|---|---|---|---|
| *P. mirabilis* | CAZ | CRO, CXM | 0.97 - 1.00 |
| *P. mirabilis* | CIP | OFX | 0.97 - 1.00 |
| *P. mirabilis* | LEX | CRO, CXM | 0.97 - 1.00 |
| *S. aureus* | CIP | OXA | 0.86 - 0.96 |
| *S. aureus* | ERY | CLI | 0.98 - 0.99 |

Note: AMC, amoxicillin/clavulanate; AMP, ampicillin; CAZ, ceftazidime; CIP, ciprofloxacin; CLI, clindamycin; CRO, ceftriaxone; CXM, cefuroxime; ERY, erythromycin; Fusid, fusidic acid; GEN, gentamicin; IPM, imipenem; LEX, cefalexin; MEM, meropenem; MUP, mupirocin; OFX, ofloxacin; OXA, oxacillin; PIP, piperacillin; SXT, sulfamethoxazole/trimethoprim, TZP, piperacillin/tazobactam.

**Table S2: The maximum number of parents sufficient to reach the maximum likelihood, the number of arcs in this model, and the number of arcs present in at least 50% of the simulations, representing the final models presented, for each of the 5 bacteria analyzed.**

| Bacteria | Site | Max parents needed | Initial # of arcs | Arcs in 50% of simulations |
|---|---|---|---|---|
| *E coli* | Urine | 7 | 42 | 39 |
| | Wound | 5 | 26 | 26 |
| | Aerobic blood | 6 | 19 | 17 |
| *K pneumoniae* | Urine | 5 | 22 | 22 |
| | Wound | 3 | 11 | 11 |
| | Aerobic blood | 3 | 9 | 9 |
| *P aeruginosa* | Urine | 6 | 19 | 18 |
| | Wound | 4 | 17 | 16 |
| | Sputum | 5 | 18 | 18 |
| *P mirabilis* | Urine | 4 | 17 | 17 |
| | Wound | 4 | 18 | 18 |
| | Aerobic blood | 2 | 7 | 4 |
| *S aureus* | Wound | 4 | 26 | 25 |
| | Aerobic blood | 2 | 8 | 7 |

*Maximum number of parents allowed

Note: Initial number of arcs is for models where variables have been removed due not being connected to the resistance variables, or in the case of Ecoli urine, variables removed to allow models to run within a reasonable (2 weeks) amount of time.

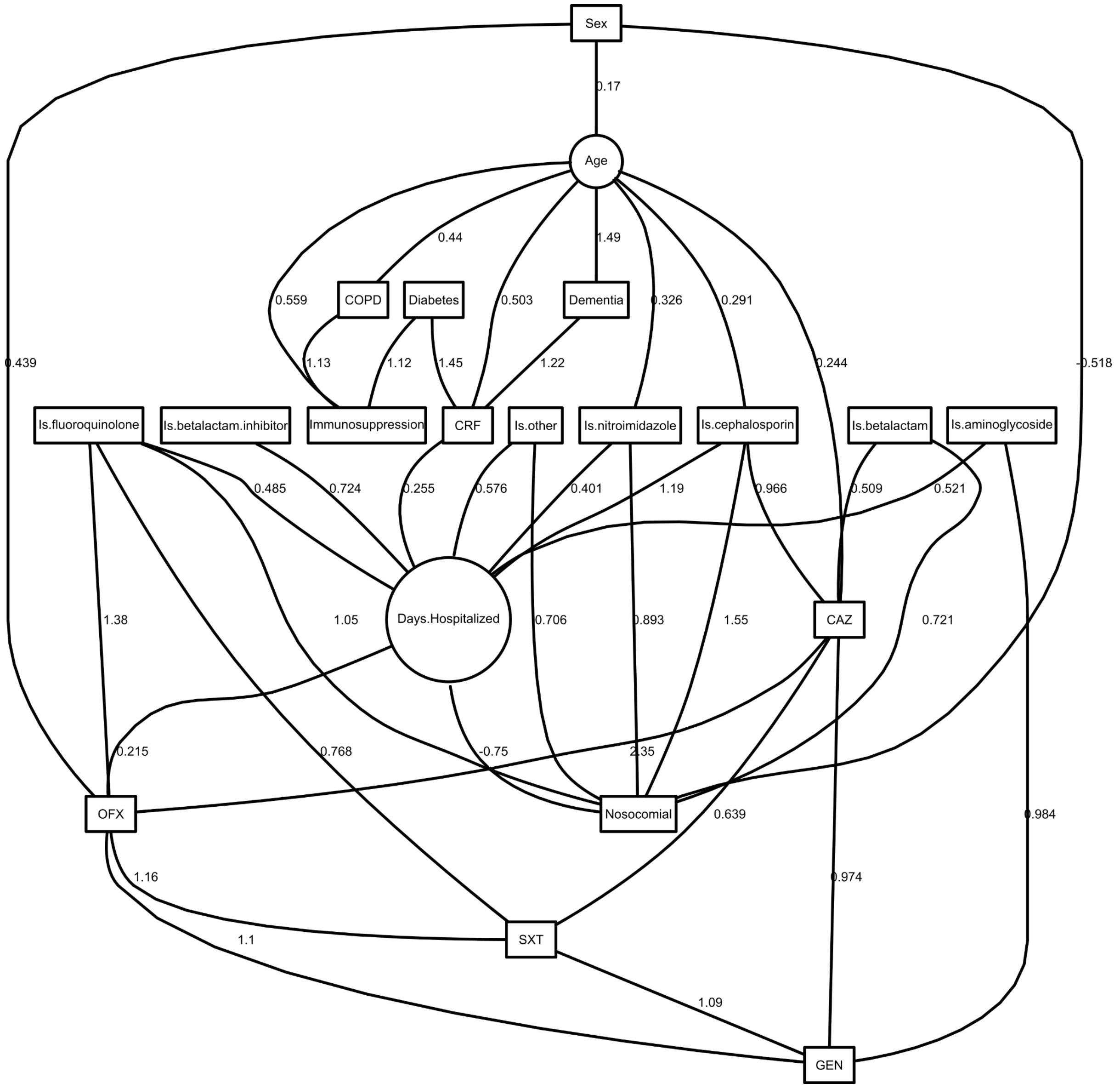

Figure S1: Final DAG for *E coli* in urine.

Table S3: *E coli* in urine parameter estimates and their 95% credible intervals. None of the non-intercept credible intervals contains zero.

| | Posterior statistics | | | |
|---|---|---|---|---|
| myvec | 2.50% | median | 97.50% | mode |
| Age\|(Intercept) | -0.10 | -0.05 | 0.00 | -0.05 |
| Age\|Sex | 0.08 | 0.17 | 0.25 | 0.17 |
| Age\|precision | 0.95 | 1.01 | 1.06 | 1.01 |
| Sex\|(Intercept) | -0.91 | -0.82 | -0.74 | -0.82 |
| Immunosuppression\|(Intercept) | -4.16 | -3.84 | -3.54 | -3.81 |
| Immunosuppression\|Age | 0.24 | 0.57 | 0.92 | 0.56 |

| | | | | |
|---|---|---|---|---|
| Immunosuppression\|Diabetes | 0.64 | 1.12 | 1.57 | 1.12 |
| Immunosuppression\|COPD | 0.40 | 1.11 | 1.74 | 1.13 |
| Dementia\|(Intercept) | -3.33 | -3.06 | -2.82 | -3.05 |
| Dementia\|Age | 1.17 | 1.50 | 1.83 | 1.49 |
| Diabetes\|(Intercept) | -1.84 | -1.72 | -1.62 | -1.72 |
| COPD\|(Intercept) | -3.41 | -3.18 | -2.98 | -3.17 |
| COPD\|Age | 0.19 | 0.44 | 0.71 | 0.44 |
| CRF\|(Intercept) | -3.85 | -3.57 | -3.31 | -3.56 |
| CRF\|Age | 0.23 | 0.51 | 0.82 | 0.50 |
| CRF\|Dementia | 0.75 | 1.22 | 1.65 | 1.22 |
| CRF\|Diabetes | 1.07 | 1.46 | 1.83 | 1.45 |
| Is.cephalosporin\|(Intercept) | -0.78 | -0.70 | -0.61 | -0.70 |
| Is.cephalosporin\|Age | 0.20 | 0.29 | 0.39 | 0.29 |
| Is.betalactam\|(Intercept) | -2.34 | -2.20 | -2.08 | -2.20 |
| Is.betalactam.inhibitor\|(Intercept) | -2.57 | -2.41 | -2.28 | -2.41 |
| Is.nitroimidazole\|(Intercept) | -2.88 | -2.70 | -2.55 | -2.70 |
| Is.nitroimidazole\|Age | 0.14 | 0.33 | 0.53 | 0.33 |
| Is.aminoglycoside\|(Intercept) | -3.11 | -2.91 | -2.74 | -2.91 |
| Is.fluoroquinolone\|(Intercept) | -3.61 | -3.38 | -3.17 | -3.38 |
| Is.other\|(Intercept) | -2.76 | -2.59 | -2.45 | -2.59 |
| Days.Hospitalized\|(Intercept) | -0.61 | -0.58 | -0.55 | -0.58 |
| Days.Hospitalized\|CRF | 0.15 | 0.25 | 0.36 | 0.26 |
| Days.Hospitalized\|Is.cephalosporin | 1.13 | 1.19 | 1.24 | 1.19 |
| Days.Hospitalized\|Is.betalactam.inhibitor | 0.63 | 0.72 | 0.81 | 0.72 |
| Days.Hospitalized\|Is.nitroimidazole | 0.29 | 0.40 | 0.51 | 0.40 |
| Days.Hospitalized\|Is.aminoglycoside | 0.40 | 0.52 | 0.63 | 0.52 |
| Days.Hospitalized\|Is.fluoroquinolone | 0.34 | 0.48 | 0.62 | 0.49 |
| Days.Hospitalized\|Is.other | 0.47 | 0.57 | 0.67 | 0.58 |
| Days.Hospitalized\|precision | 2.49 | 2.64 | 2.79 | 2.65 |
| Nosocomial\|(Intercept) | -2.18 | -1.99 | -1.82 | -1.99 |
| Nosocomial\|Sex | -0.77 | -0.52 | -0.29 | -0.52 |
| Nosocomial\|Is.cephalosporin | 1.26 | 1.57 | 1.86 | 1.55 |
| Nosocomial\|Is.betalactam | 0.36 | 0.73 | 1.06 | 0.72 |
| Nosocomial\|Is.nitroimidazole | 0.50 | 0.90 | 1.29 | 0.89 |
| Nosocomial\|Is.fluoroquinolone | 0.51 | 1.06 | 1.59 | 1.05 |
| Nosocomial\|Is.other | 0.32 | 0.71 | 1.07 | 0.71 |
| Nosocomial\|Days.Hospitalized | -0.94 | -0.76 | -0.59 | -0.75 |

| | | | | |
|---|---|---|---|---|
| CAZ\|(Intercept) | -1.22 | -1.10 | -0.99 | -1.10 |
| CAZ\|Age | 0.15 | 0.24 | 0.34 | 0.24 |
| CAZ\|Is.cephalosporin | 0.78 | 0.97 | 1.15 | 0.97 |
| CAZ\|Is.betalactam | 0.22 | 0.52 | 0.79 | 0.51 |
| GEN\|(Intercept) | -3.65 | -3.39 | -3.13 | -3.37 |
| GEN\|Is.aminoglycoside | 0.53 | 0.99 | 1.42 | 0.98 |
| GEN\|CAZ | 0.70 | 0.98 | 1.25 | 0.97 |
| GEN\|OFX | 0.81 | 1.10 | 1.38 | 1.10 |
| GEN\|SXT | 0.83 | 1.10 | 1.35 | 1.09 |
| OFX\|(Intercept) | -1.96 | -1.80 | -1.65 | -1.80 |
| OFX\|Sex | 0.22 | 0.44 | 0.65 | 0.44 |
| OFX\|Is.fluoroquinolone | 0.83 | 1.38 | 1.96 | 1.38 |
| OFX\|Days.Hospitalized | 0.11 | 0.22 | 0.31 | 0.21 |
| OFX\|CAZ | 2.15 | 2.35 | 2.56 | 2.35 |
| SXT\|(Intercept) | -1.48 | -1.35 | -1.22 | -1.35 |
| SXT\|Is.fluoroquinolone | 0.27 | 0.78 | 1.26 | 0.77 |
| SXT\|CAZ | 0.43 | 0.64 | 0.84 | 0.64 |
| SXT\|OFX | 0.96 | 1.17 | 1.37 | 1.16 |
| | OR | | | |
| | 2.50% | median | 97.50% | mode |
| Age\|Sex | 1.08 | 1.18 | 1.29 | 1.18 |
| Immunosuppression\|Age | 1.27 | 1.76 | 2.50 | 1.75 |
| Immunosuppression\|Diabetes | 1.89 | 3.07 | 4.82 | 3.07 |
| Immunosuppression\|COPD | 1.49 | 3.03 | 5.67 | 3.08 |
| Dementia\|Age | 3.23 | 4.50 | 6.26 | 4.46 |
| COPD\|Age | 1.21 | 1.56 | 2.04 | 1.55 |
| CRF\|Age | 1.26 | 1.67 | 2.26 | 1.65 |
| CRF\|Dementia | 2.12 | 3.37 | 5.18 | 3.39 |
| CRF\|Diabetes | 2.92 | 4.29 | 6.22 | 4.26 |
| Is.cephalosporin\|Age | 1.22 | 1.34 | 1.47 | 1.34 |
| Is.nitroimidazole\|Age | 1.15 | 1.39 | 1.71 | 1.39 |
| Nosocomial\|Sex | 0.46 | 0.60 | 0.75 | 0.60 |
| Nosocomial\|Is.cephalosporin | 3.53 | 4.78 | 6.40 | 4.72 |
| Nosocomial\|Is.betalactam | 1.44 | 2.07 | 2.90 | 2.06 |
| Nosocomial\|Is.nitroimidazole | 1.64 | 2.46 | 3.64 | 2.44 |
| Nosocomial\|Is.fluoroquinolone | 1.67 | 2.89 | 4.92 | 2.86 |

| | | | | |
|---|---|---|---|---|
| Nosocomial\|Is.other | 1.38 | 2.04 | 2.93 | 2.03 |
| Nosocomial\|Days.Hospitalized | 0.39 | 0.47 | 0.55 | 0.47 |
| CAZ\|Age | 1.16 | 1.28 | 1.40 | 1.28 |
| CAZ\|Is.cephalosporin | 2.19 | 2.64 | 3.15 | 2.63 |
| CAZ\|Is.betalactam | 1.24 | 1.67 | 2.20 | 1.66 |
| GEN\|Is.aminoglycoside | 1.69 | 2.70 | 4.14 | 2.67 |
| GEN\|CAZ | 2.01 | 2.66 | 3.50 | 2.65 |
| GEN\|OFX | 2.25 | 3.02 | 3.99 | 3.00 |
| GEN\|SXT | 2.30 | 3.00 | 3.87 | 2.99 |
| OFX\|Sex | 1.25 | 1.56 | 1.91 | 1.55 |
| OFX\|Is.fluoroquinolone | 2.29 | 3.99 | 7.10 | 3.99 |
| OFX\|Days.Hospitalized | 1.12 | 1.24 | 1.37 | 1.24 |
| OFX\|CAZ | 8.61 | 10.50 | 12.90 | 10.48 |
| SXT\|Is.fluoroquinolone | 1.31 | 2.18 | 3.53 | 2.16 |
| SXT\|CAZ | 1.53 | 1.90 | 2.32 | 1.90 |
| SXT\|OFX | 2.60 | 3.21 | 3.94 | 3.20 |

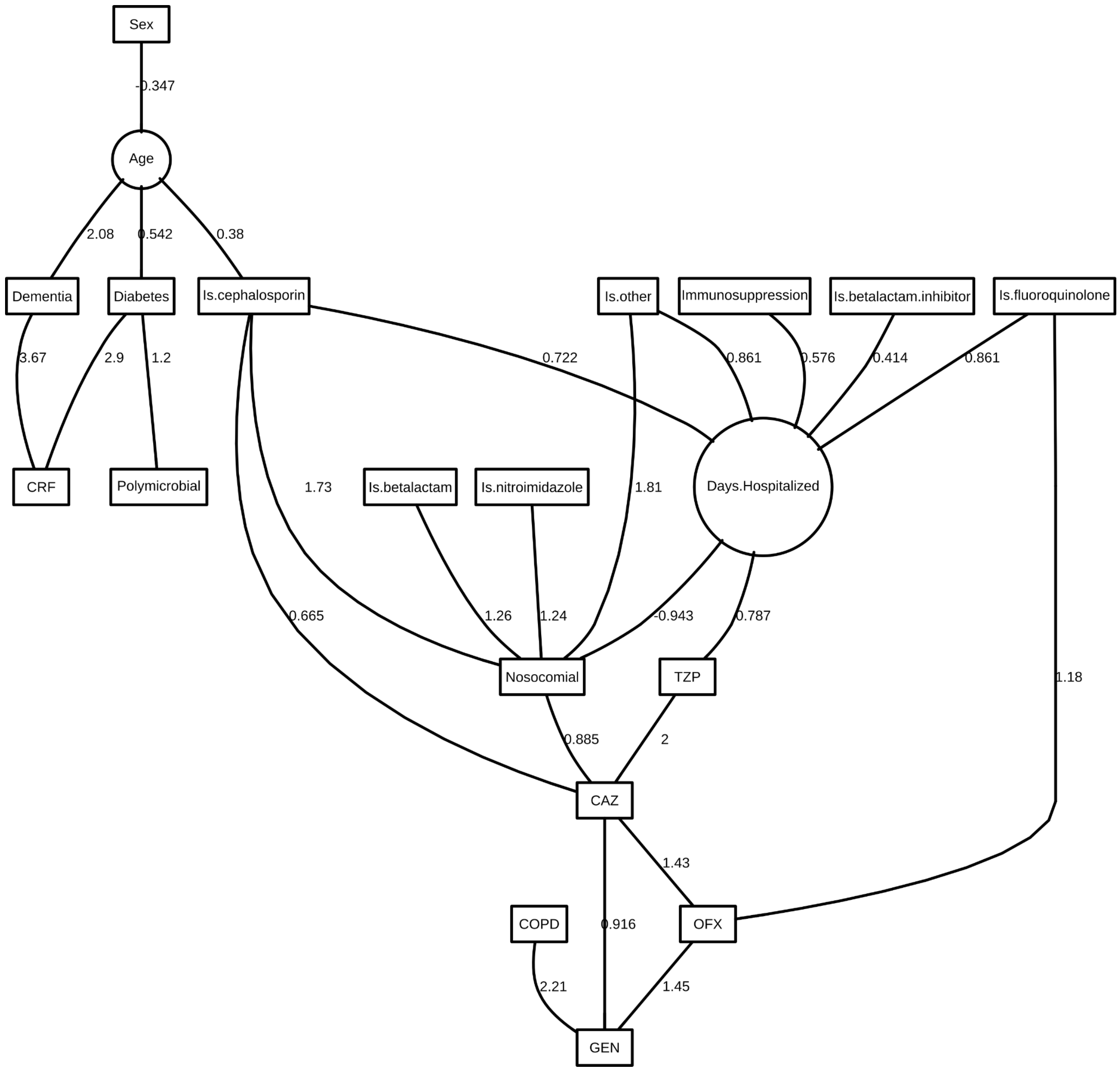

Figure S2: Final DAG for *E coli* in wound.

Table S4: *E coli* in wound parameter estimates and their 95% credible intervals. None of the non-intercept credible intervals contains zero.

| | Posterior statistics | | | |
|---|---|---|---|---|
| myvec | 2.50% | median | 97.50% | mode |
| Age\|(Intercept) | 0.07 | 0.20 | 0.32 | 0.20 |
| Age\|Sex | -0.51 | -0.34 | -0.19 | -0.35 |
| Age\|precision | 0.91 | 1.03 | 1.15 | 1.03 |
| Sex\|(Intercept) | 0.14 | 0.31 | 0.47 | 0.31 |
| Immunosuppression\|(Intercept) | -3.70 | -3.23 | -2.84 | -3.22 |
| Dementia\|(Intercept) | -7.59 | -5.67 | -4.39 | -5.49 |
| Dementia\|Age | 1.03 | 2.18 | 3.58 | 2.08 |

| | | | | |
|---|---|---|---|---|
| Diabetes\|(Intercept) | -2.00 | -1.74 | -1.51 | -1.73 |
| Diabetes\|Age | 0.29 | 0.55 | 0.80 | 0.54 |
| COPD\|(Intercept) | -4.35 | -3.74 | -3.25 | -3.73 |
| CRF\|(Intercept) | -6.21 | -4.92 | -4.05 | -4.84 |
| CRF\|Dementia | 1.71 | 3.71 | 5.49 | 3.67 |
| CRF\|Diabetes | 1.84 | 3.00 | 4.31 | 2.90 |
| Is.cephalosporin\|(Intercept) | -0.06 | 0.11 | 0.28 | 0.11 |
| Is.cephalosporin\|Age | 0.21 | 0.38 | 0.55 | 0.38 |
| Is.betalactam\|(Intercept) | -0.93 | -0.74 | -0.57 | -0.74 |
| Is.betalactam.inhibitor\|(Intercept) | -1.62 | -1.40 | -1.20 | -1.40 |
| Is.nitroimidazole\|(Intercept) | -0.89 | -0.70 | -0.54 | -0.71 |
| Is.fluoroquinolone\|(Intercept) | -2.99 | -2.64 | -2.34 | -2.63 |
| Is.other\|(Intercept) | -2.56 | -2.28 | -2.00 | -2.27 |
| Days.Hospitalized\|(Intercept) | -0.73 | -0.62 | -0.52 | -0.62 |
| Days.Hospitalized\|Immunosuppression | 0.22 | 0.57 | 0.92 | 0.58 |
| Days.Hospitalized\|Is.cephalosporin | 0.58 | 0.72 | 0.85 | 0.72 |
| Days.Hospitalized\|Is.betalactam.inhibitor | 0.24 | 0.41 | 0.58 | 0.41 |
| Days.Hospitalized\|Is.fluoroquinolone | 0.58 | 0.86 | 1.13 | 0.86 |
| Days.Hospitalized\|Is.other | 0.63 | 0.86 | 1.09 | 0.86 |
| Days.Hospitalized\|precision | 1.37 | 1.55 | 1.73 | 1.56 |
| Nosocomial\|(Intercept) | -1.56 | -1.22 | -0.88 | -1.19 |
| Nosocomial\|Is.cephalosporin | 1.24 | 1.77 | 2.27 | 1.73 |
| Nosocomial\|Is.betalactam | 0.80 | 1.28 | 1.75 | 1.26 |
| Nosocomial\|Is.nitroimidazole | 0.72 | 1.25 | 1.80 | 1.24 |
| Nosocomial\|Is.other | 0.99 | 1.86 | 2.78 | 1.81 |
| Nosocomial\|Days.Hospitalized | -1.26 | -0.96 | -0.70 | -0.94 |
| Polymicrobial\|(Intercept) | 0.57 | 0.76 | 0.95 | 0.76 |
| Polymicrobial\|Diabetes | 0.59 | 1.22 | 1.92 | 1.20 |
| CAZ\|(Intercept) | -2.45 | -2.02 | -1.62 | -2.00 |
| CAZ\|Is.cephalosporin | 0.25 | 0.67 | 1.08 | 0.66 |
| CAZ\|Nosocomial | 0.45 | 0.90 | 1.35 | 0.89 |
| CAZ\|TZP | 1.05 | 2.06 | 3.12 | 2.00 |
| GEN\|(Intercept) | -2.90 | -2.51 | -2.16 | -2.50 |
| GEN\|COPD | 1.01 | 2.23 | 3.41 | 2.21 |
| GEN\|CAZ | 0.42 | 0.93 | 1.42 | 0.92 |
| GEN\|OFX | 0.96 | 1.46 | 1.95 | 1.45 |
| OFX\|(Intercept) | -2.00 | -1.71 | -1.45 | -1.70 |

| | | | | |
|---|---|---|---|---|
| OFX\|Is.fluoroquinolone | 0.45 | 1.20 | 1.89 | 1.18 |
| OFX\|CAZ | 1.03 | 1.44 | 1.84 | 1.43 |
| TZP\|(Intercept) | -4.26 | -3.63 | -3.13 | -3.60 |
| TZP\|Days.Hospitalized | 0.40 | 0.80 | 1.19 | 0.79 |
| | OR | | | |
| | 2.50% | median | 97.50% | mode |
| Age\|Sex | 0.60 | 0.71 | 0.83 | 0.71 |
| Dementia\|Age | 2.79 | 8.88 | 35.97 | 7.97 |
| Diabetes\|Age | 1.34 | 1.73 | 2.23 | 1.72 |
| CRF\|Dementia | 5.51 | 40.89 | 242.73 | 39.40 |
| CRF\|Diabetes | 6.29 | 20.08 | 74.17 | 18.23 |
| Is.cephalosporin\|Age | 1.23 | 1.46 | 1.73 | 1.46 |
| Nosocomial\|Is.cephalosporin | 3.46 | 5.84 | 9.67 | 5.65 |
| Nosocomial\|Is.betalactam | 2.22 | 3.58 | 5.77 | 3.53 |
| Nosocomial\|Is.nitroimidazole | 2.05 | 3.49 | 6.06 | 3.46 |
| Nosocomial\|Is.other | 2.68 | 6.44 | 16.06 | 6.11 |
| Nosocomial\|Days.Hospitalized | 0.28 | 0.38 | 0.50 | 0.39 |
| Polymicrobial\|Diabetes | 1.80 | 3.40 | 6.79 | 3.31 |
| CAZ\|Is.cephalosporin | 1.29 | 1.96 | 2.93 | 1.94 |
| CAZ\|Nosocomial | 1.57 | 2.46 | 3.86 | 2.42 |
| CAZ\|TZP | 2.85 | 7.86 | 22.64 | 7.35 |
| GEN\|COPD | 2.73 | 9.31 | 30.21 | 9.09 |
| GEN\|CAZ | 1.52 | 2.53 | 4.12 | 2.50 |
| GEN\|OFX | 2.62 | 4.29 | 7.02 | 4.26 |
| OFX\|Is.fluoroquinolone | 1.57 | 3.31 | 6.60 | 3.27 |
| OFX\|CAZ | 2.79 | 4.23 | 6.31 | 4.19 |
| TZP\|Days.Hospitalized | 1.49 | 2.23 | 3.27 | 2.20 |

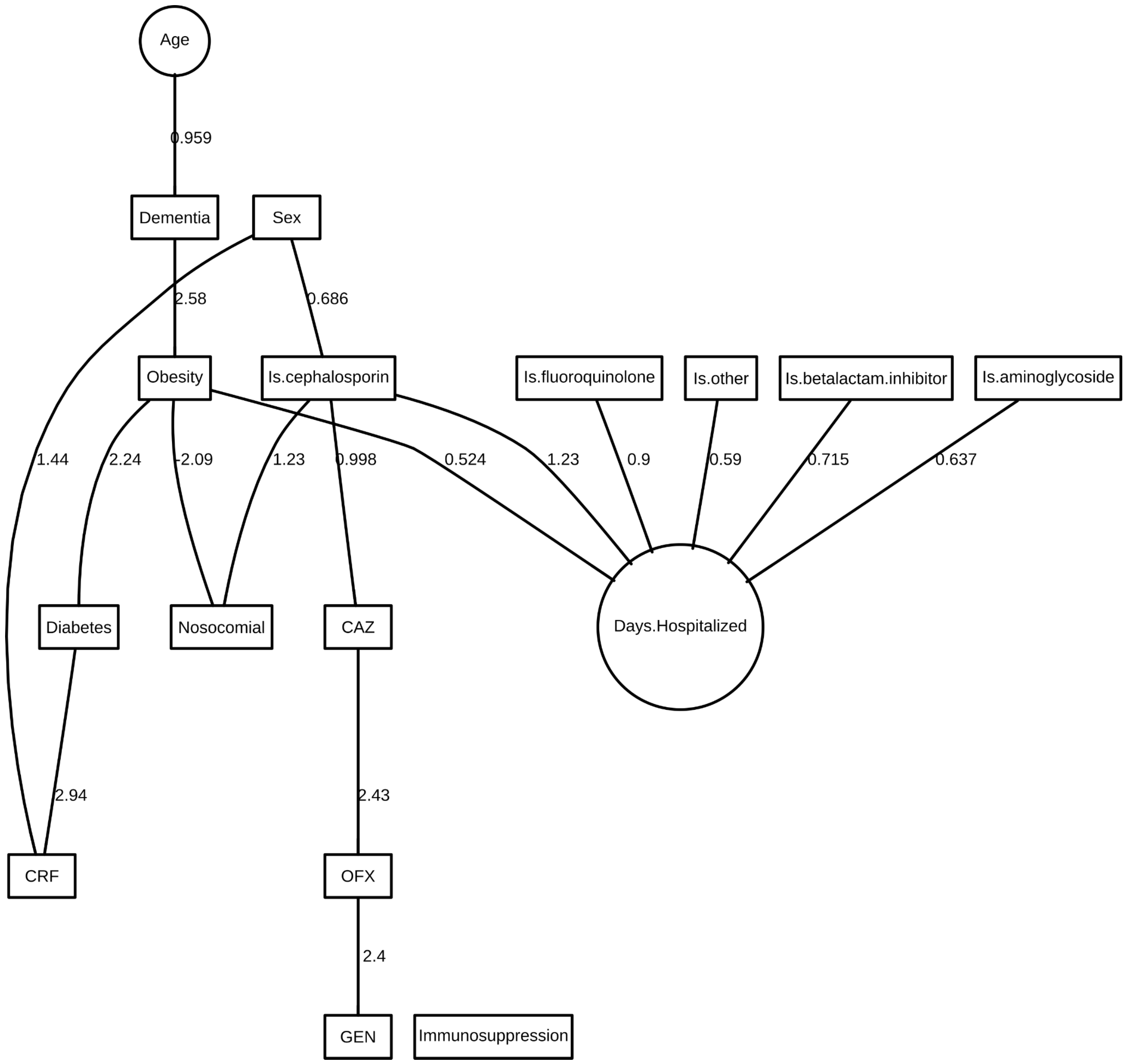

Figure S3: Final DAG for *E coli* in aerobic blood.

Table S5: *E coli* in aerobic blood parameter estimates and their 95% credible intervals. None of the non-intercept credible intervals contains zero.

| | Posterior statistics | | | |
|---|---|---|---|---|
| myvec | 2.50% | median | 97.50% | mode |
| Age\|(Intercept) | -0.11 | 0.00 | 0.10 | 0.00 |
| Age\|precision | 0.86 | 1.01 | 1.15 | 1.00 |
| Sex\|(Intercept) | -0.65 | -0.43 | -0.22 | -0.43 |
| Immunosuppression\|(Intercept) | -3.17 | -2.71 | -2.32 | -2.71 |
| Dementia\|(Intercept) | -3.76 | -3.09 | -2.59 | -3.05 |
| Dementia\|Age | 0.35 | 1.00 | 1.73 | 0.96 |
| Diabetes\|(Intercept) | -2.21 | -1.88 | -1.58 | -1.87 |

| | | | | |
|---|---|---|---|---|
| Diabetes\|Obesity | 1.31 | 2.28 | 3.21 | 2.24 |
| CRF\|(Intercept) | -6.18 | -4.85 | -3.88 | -4.74 |
| CRF\|Sex | 0.45 | 1.51 | 2.60 | 1.44 |
| CRF\|Diabetes | 2.01 | 3.01 | 4.09 | 2.94 |
| Obesity\|(Intercept) | -3.83 | -3.22 | -2.72 | -3.21 |
| Obesity\|Dementia | 1.48 | 2.57 | 3.62 | 2.58 |
| Is.cephalosporin\|(Intercept) | -0.99 | -0.70 | -0.43 | -0.70 |
| Is.cephalosporin\|Sex | 0.24 | 0.70 | 1.11 | 0.69 |
| Is.betalactam.inhibitor\|(Intercept) | -2.73 | -2.33 | -2.00 | -2.33 |
| Is.aminoglycoside\|(Intercept) | -3.64 | -3.09 | -2.63 | -3.07 |
| Is.fluoroquinolone\|(Intercept) | -4.94 | -4.04 | -3.35 | -3.98 |
| Is.other\|(Intercept) | -3.30 | -2.81 | -2.40 | -2.80 |
| Days.Hospitalized\|(Intercept) | -0.74 | -0.66 | -0.58 | -0.66 |
| Days.Hospitalized\|Obesity | 0.26 | 0.52 | 0.77 | 0.52 |
| Days.Hospitalized\|Is.cephalosporin | 1.09 | 1.22 | 1.36 | 1.23 |
| Days.Hospitalized\|Is.betalactam.inhibitor | 0.48 | 0.72 | 0.94 | 0.71 |
| Days.Hospitalized\|Is.aminoglycoside | 0.33 | 0.63 | 0.94 | 0.64 |
| Days.Hospitalized\|Is.fluoroquinolone | 0.43 | 0.89 | 1.35 | 0.90 |
| Days.Hospitalized\|Is.other | 0.31 | 0.58 | 0.86 | 0.59 |
| Days.Hospitalized\|precision | 2.50 | 2.93 | 3.36 | 2.97 |
| Nosocomial\|(Intercept) | -2.20 | -1.80 | -1.43 | -1.78 |
| Nosocomial\|Obesity | -5.85 | -2.41 | -0.68 | -2.09 |
| Nosocomial\|Is.cephalosporin | 0.72 | 1.23 | 1.77 | 1.23 |
| CAZ\|(Intercept) | -1.97 | -1.60 | -1.26 | -1.59 |
| CAZ\|Is.cephalosporin | 0.50 | 1.01 | 1.49 | 1.00 |
| GEN\|(Intercept) | -4.10 | -3.35 | -2.78 | -3.32 |
| GEN\|OFX | 1.65 | 2.44 | 3.26 | 2.40 |
| OFX\|(Intercept) | -2.31 | -1.93 | -1.60 | -1.92 |
| OFX\|CAZ | 1.88 | 2.43 | 3.01 | 2.43 |
| | OR | | | |
| | 2.50% | median | 97.50% | mode |
| Dementia\|Age | 1.42 | 2.72 | 5.66 | 2.61 |
| Diabetes\|Obesity | 3.70 | 9.77 | 24.87 | 9.41 |
| CRF\|Sex | 1.57 | 4.52 | 13.51 | 4.22 |
| CRF\|Diabetes | 7.45 | 20.23 | 59.68 | 18.95 |
| Obesity\|Dementia | 4.41 | 13.12 | 37.46 | 13.17 |

| | | | | |
|---|---|---|---|---|
| Is.cephalosporin\|Sex | 1.28 | 2.00 | 3.03 | 1.99 |
| Nosocomial\|Obesity | 0.00 | 0.09 | 0.50 | 0.12 |
| Nosocomial\|Is.cephalosporin | 2.05 | 3.43 | 5.85 | 3.43 |
| CAZ\|Is.cephalosporin | 1.66 | 2.74 | 4.44 | 2.71 |
| GEN\|OFX | 5.23 | 11.49 | 26.09 | 11.03 |
| OFX\|CAZ | 6.56 | 11.39 | 20.22 | 11.33 |

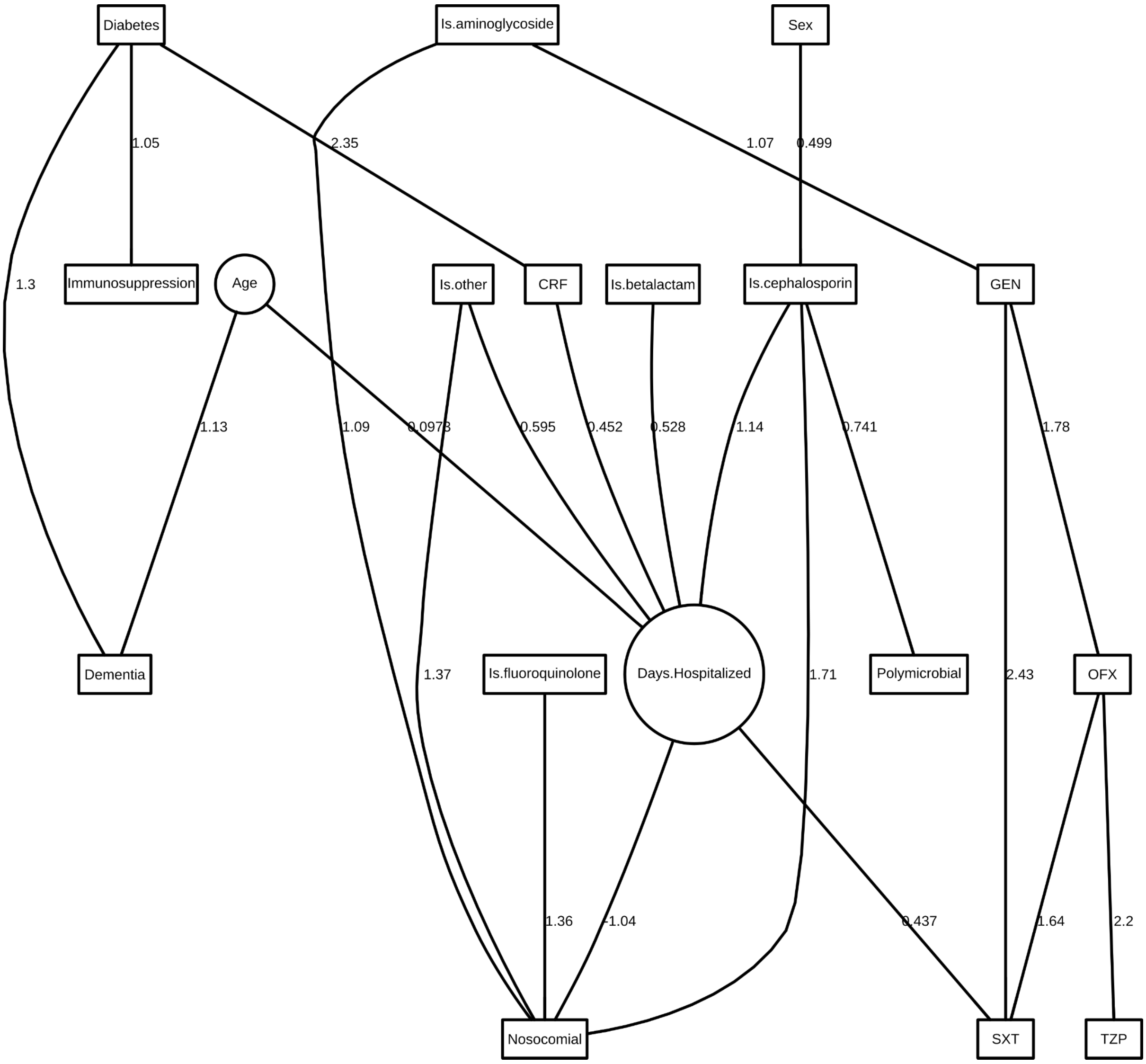

Figure S4: Final DAG for *K pneumoniae* in urine.

Table S6: *K pneumoniae* in urine parameter estimates and their 95% credible intervals. None of the non-intercept credible intervals contains zero.

| | Posterior statistics | | | |
|---|---|---|---|---|
| myvec | 2.50% | median | 97.50% | mode |

| | | | | |
|---|---|---|---|---|
| Age\|(Intercept) | -0.07 | 0.00 | 0.07 | 0.00 |
| Age\|precision | 0.90 | 1.00 | 1.10 | 1.00 |
| Sex\|(Intercept) | -0.48 | -0.33 | -0.20 | -0.33 |
| Immunosuppression\|(Intercept) | -3.61 | -3.17 | -2.80 | -3.16 |
| Immunosuppression\|Diabetes | 0.38 | 1.04 | 1.68 | 1.05 |
| Dementia\|(Intercept) | -3.65 | -3.17 | -2.75 | -3.14 |
| Dementia\|Age | 0.67 | 1.15 | 1.66 | 1.13 |
| Dementia\|Diabetes | 0.72 | 1.31 | 1.88 | 1.30 |
| Diabetes\|(Intercept) | -1.59 | -1.41 | -1.24 | -1.41 |
| CRF\|(Intercept) | -4.17 | -3.62 | -3.16 | -3.60 |
| CRF\|Diabetes | 1.75 | 2.38 | 3.00 | 2.35 |
| Is.cephalosporin\|(Intercept) | -0.56 | -0.36 | -0.18 | -0.36 |
| Is.cephalosporin\|Sex | 0.20 | 0.51 | 0.78 | 0.50 |
| Is.betalactam\|(Intercept) | -1.85 | -1.65 | -1.47 | -1.65 |
| Is.aminoglycoside\|(Intercept) | -2.63 | -2.36 | -2.12 | -2.36 |
| Is.fluoroquinolone\|(Intercept) | -3.30 | -2.96 | -2.65 | -2.94 |
| Is.other\|(Intercept) | -2.46 | -2.21 | -1.99 | -2.21 |
| Days.Hospitalized\|(Intercept) | -0.77 | -0.70 | -0.63 | -0.70 |
| Days.Hospitalized\|Age | 0.05 | 0.10 | 0.15 | 0.10 |
| Days.Hospitalized\|CRF | 0.25 | 0.45 | 0.65 | 0.45 |
| Days.Hospitalized\|Is.cephalosporin | 1.03 | 1.13 | 1.24 | 1.14 |
| Days.Hospitalized\|Is.betalactam | 0.38 | 0.52 | 0.66 | 0.53 |
| Days.Hospitalized\|Is.other | 0.43 | 0.59 | 0.76 | 0.60 |
| Days.Hospitalized\|precision | 1.88 | 2.09 | 2.30 | 2.10 |
| Nosocomial\|(Intercept) | -2.34 | -2.01 | -1.71 | -2.00 |
| Nosocomial\|Is.cephalosporin | 1.27 | 1.73 | 2.18 | 1.71 |
| Nosocomial\|Is.aminoglycoside | 0.48 | 1.11 | 1.72 | 1.09 |
| Nosocomial\|Is.fluoroquinolone | 0.59 | 1.38 | 2.17 | 1.36 |
| Nosocomial\|Is.other | 0.79 | 1.38 | 1.96 | 1.37 |
| Nosocomial\|Days.Hospitalized | -1.35 | -1.06 | -0.80 | -1.04 |
| Polymicrobial\|(Intercept) | -2.35 | -2.03 | -1.75 | -2.03 |
| Polymicrobial\|Is.cephalosporin | 0.34 | 0.75 | 1.13 | 0.74 |
| GEN\|(Intercept) | -1.65 | -1.46 | -1.27 | -1.45 |
| GEN\|Is.aminoglycoside | 0.52 | 1.06 | 1.58 | 1.07 |
| OFX\|(Intercept) | -2.91 | -2.57 | -2.29 | -2.57 |
| OFX\|GEN | 1.33 | 1.78 | 2.23 | 1.78 |
| SXT\|(Intercept) | -1.20 | -1.01 | -0.83 | -1.01 |

| | | | | |
|---|---|---|---|---|
| SXT\|Days.Hospitalized | 0.27 | 0.44 | 0.60 | 0.44 |
| SXT\|GEN | 1.97 | 2.44 | 2.96 | 2.43 |
| SXT\|OFX | 1.07 | 1.64 | 2.27 | 1.64 |
| TZP\|(Intercept) | -4.42 | -3.84 | -3.38 | -3.82 |
| TZP\|OFX | 1.44 | 2.22 | 2.96 | 2.20 |
| | OR | | | |
| | 2.50% | median | 97.50% | mode |
| Immunosuppression\|Diabetes | 1.46 | 2.84 | 5.37 | 2.87 |
| Dementia\|Age | 1.95 | 3.17 | 5.25 | 3.09 |
| Dementia\|Diabetes | 2.06 | 3.71 | 6.54 | 3.67 |
| CRF\|Diabetes | 5.76 | 10.75 | 20.07 | 10.53 |
| Is.cephalosporin\|Sex | 1.22 | 1.66 | 2.19 | 1.65 |
| Nosocomial\|Is.cephalosporin | 3.58 | 5.63 | 8.87 | 5.55 |
| Nosocomial\|Is.aminoglycoside | 1.61 | 3.04 | 5.59 | 2.99 |
| Nosocomial\|Is.fluoroquinolone | 1.81 | 3.98 | 8.78 | 3.90 |
| Nosocomial\|Is.other | 2.20 | 3.96 | 7.12 | 3.92 |
| Nosocomial\|Days.Hospitalized | 0.26 | 0.35 | 0.45 | 0.35 |
| Polymicrobial\|Is.cephalosporin | 1.40 | 2.12 | 3.09 | 2.10 |
| GEN\|Is.aminoglycoside | 1.68 | 2.89 | 4.87 | 2.92 |
| OFX\|GEN | 3.77 | 5.93 | 9.34 | 5.95 |
| SXT\|Days.Hospitalized | 1.31 | 1.55 | 1.83 | 1.55 |
| SXT\|GEN | 7.19 | 11.53 | 19.22 | 11.40 |
| SXT\|OFX | 2.91 | 5.17 | 9.64 | 5.14 |
| TZP\|OFX | 4.21 | 9.17 | 19.35 | 9.03 |

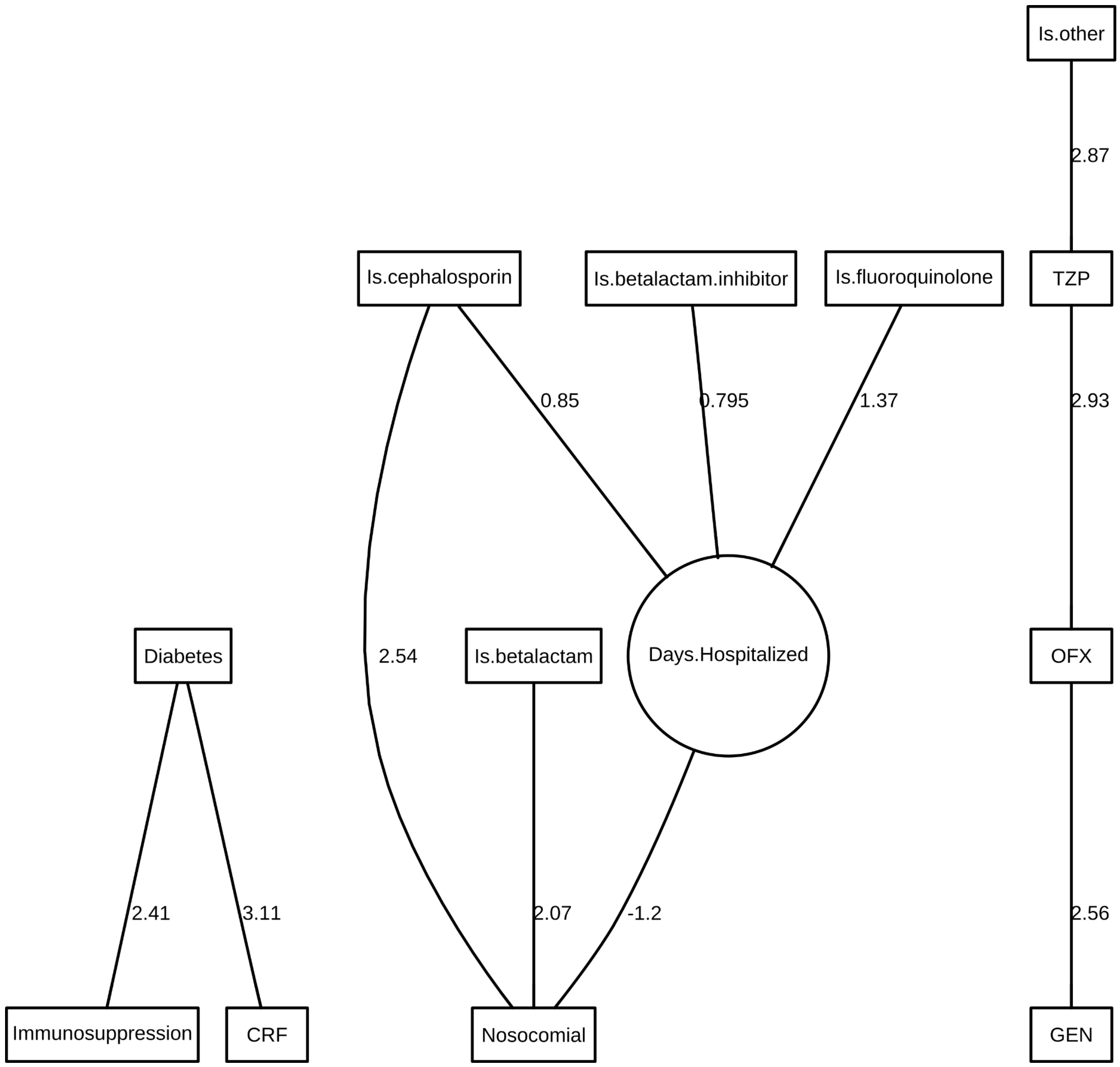

Figure S5: Final DAG for *K pneumoniae* in wound.

Table S7: *K pneumoniae* in wound parameter estimates and their 95% credible intervals. None of the non-intercept credible intervals contains zero.

| | Posterior statistics | | | |
|---|---|---|---|---|
| myvec | 2.50% | median | 97.50% | mode |
| Immunosuppression\|(Intercept) | -6.56 | -4.58 | -3.34 | -4.38 |
| Immunosuppression\|Diabetes | 0.96 | 2.51 | 4.56 | 2.41 |
| Diabetes\|(Intercept) | -1.37 | -1.05 | -0.75 | -1.04 |
| CRF\|(Intercept) | -5.59 | -4.09 | -3.08 | -3.97 |
| CRF\|Diabetes | 2.00 | 3.23 | 4.78 | 3.11 |
| Is.cephalosporin\|(Intercept) | 0.28 | 0.57 | 0.85 | 0.57 |
| Is.betalactam\|(Intercept) | -0.84 | -0.55 | -0.28 | -0.55 |

| | | | | |
|---|---|---|---|---|
| Is.betalactam.inhibitor\|(Intercept) | -1.37 | -1.05 | -0.75 | -1.04 |
| Is.fluoroquinolone\|(Intercept) | -3.33 | -2.71 | -2.19 | -2.68 |
| Is.other\|(Intercept) | -2.32 | -1.89 | -1.51 | -1.88 |
| Days.Hospitalized\|(Intercept) | -1.02 | -0.83 | -0.66 | -0.84 |
| Days.Hospitalized\|Is.cephalosporin | 0.63 | 0.85 | 1.06 | 0.85 |
| Days.Hospitalized\|Is.betalactam.inhibitor | 0.56 | 0.79 | 1.02 | 0.80 |
| Days.Hospitalized\|Is.fluoroquinolone | 0.95 | 1.37 | 1.78 | 1.37 |
| Days.Hospitalized\|precision | 1.46 | 1.79 | 2.14 | 1.81 |
| Nosocomial\|(Intercept) | -2.15 | -1.43 | -0.78 | -1.40 |
| Nosocomial\|Is.cephalosporin | 1.74 | 2.60 | 3.53 | 2.54 |
| Nosocomial\|Is.betalactam | 1.27 | 2.12 | 3.05 | 2.07 |
| Nosocomial\|Days.Hospitalized | -1.77 | -1.24 | -0.76 | -1.20 |
| GEN\|(Intercept) | -2.07 | -1.65 | -1.27 | -1.64 |
| GEN\|OFX | 1.68 | 2.58 | 3.55 | 2.56 |
| OFX\|(Intercept) | -2.76 | -2.24 | -1.82 | -2.24 |
| OFX\|TZP | 1.65 | 2.98 | 4.42 | 2.93 |
| TZP\|(Intercept) | -5.20 | -3.92 | -3.03 | -3.84 |
| TZP\|Is.other | 1.62 | 2.92 | 4.39 | 2.87 |
| | OR | | | |
| | 2.50% | median | 97.50% | mode |
| Immunosuppression\|Diabetes | 2.61 | 12.36 | 95.92 | 11.16 |
| CRF\|Diabetes | 7.37 | 25.20 | 118.68 | 22.47 |
| Nosocomial\|Is.cephalosporin | 5.68 | 13.46 | 34.27 | 12.72 |
| Nosocomial\|Is.betalactam | 3.56 | 8.37 | 21.14 | 7.96 |
| Nosocomial\|Days.Hospitalized | 0.17 | 0.29 | 0.47 | 0.30 |
| GEN\|OFX | 5.37 | 13.20 | 34.95 | 12.90 |
| OFX\|TZP | 5.21 | 19.69 | 82.87 | 18.67 |
| TZP\|Is.other | 5.05 | 18.62 | 80.36 | 17.68 |

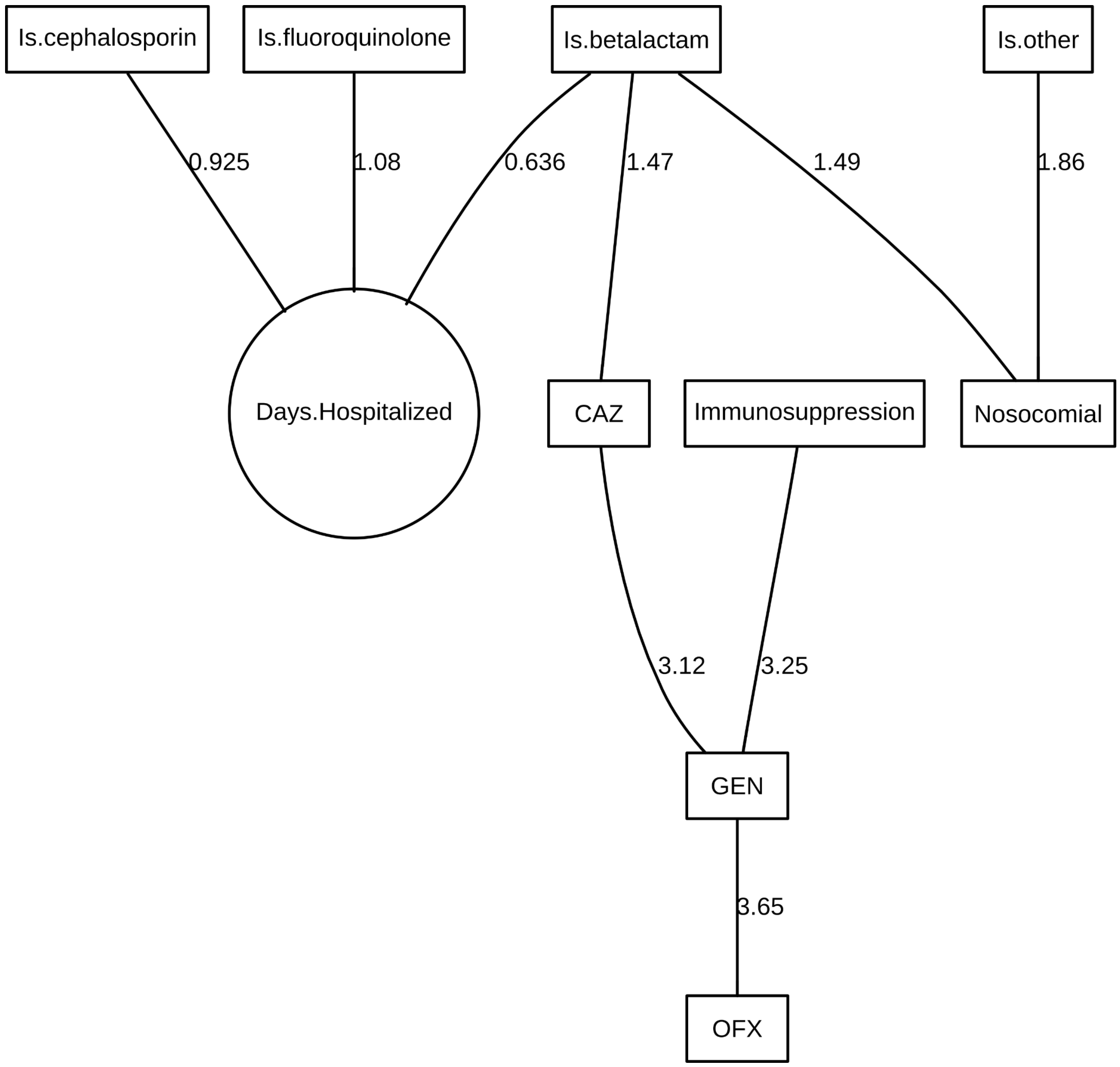

Figure S6: Final DAG for *K pneumoniae* in aerobic blood.

Table S8: *K pneumoniae* in aerobic blood parameter estimates and their 95% credible intervals. None of the non-intercept credible intervals contains zero.

| | Posterior statistics | | | |
|---|---|---|---|---|
| myvec | 2.50% | median | 97.50% | mode |
| Immunosuppression\|(Intercept) | -3.88 | -2.88 | -2.13 | -2.83 |
| Is.cephalosporin\|(Intercept) | -0.20 | 0.19 | 0.57 | 0.19 |
| Is.betalactam\|(Intercept) | -1.76 | -1.27 | -0.83 | -1.25 |
| Is.fluoroquinolone\|(Intercept) | -3.88 | -2.88 | -2.13 | -2.83 |
| Is.other\|(Intercept) | -2.36 | -1.76 | -1.27 | -1.75 |
| Days.Hospitalized\|(Intercept) | -0.92 | -0.70 | -0.50 | -0.71 |
| Days.Hospitalized\|Is.cephalosporin | 0.62 | 0.92 | 1.22 | 0.92 |

| | | | | |
|---|---|---|---|---|
| Days.Hospitalized\|Is.betalactam | 0.27 | 0.63 | 0.98 | 0.64 |
| Days.Hospitalized\|Is.fluoroquinolone | 0.41 | 1.07 | 1.72 | 1.08 |
| Days.Hospitalized\|precision | 1.36 | 1.81 | 2.37 | 1.87 |
| Nosocomial\|(Intercept) | -1.53 | -0.97 | -0.47 | -0.95 |
| Nosocomial\|Is.betalactam | 0.52 | 1.54 | 2.60 | 1.49 |
| Nosocomial\|Is.other | 0.68 | 1.94 | 3.36 | 1.86 |
| CAZ\|(Intercept) | -1.88 | -1.32 | -0.80 | -1.30 |
| CAZ\|Is.betalactam | 0.48 | 1.48 | 2.48 | 1.47 |
| GEN\|(Intercept) | -4.81 | -3.32 | -2.27 | -3.17 |
| GEN\|Immunosuppression | 1.19 | 3.49 | 6.06 | 3.25 |
| GEN\|CAZ | 1.90 | 3.26 | 4.84 | 3.12 |
| OFX\|(Intercept) | -5.93 | -3.93 | -2.68 | -3.75 |
| OFX\|GEN | 2.22 | 3.83 | 5.93 | 3.65 |
| | OR | | | |
| | 2.50% | median | 97.50% | mode |
| Nosocomial\|Is.betalactam | 1.69 | 4.67 | 13.42 | 4.44 |
| Nosocomial\|Is.other | 1.97 | 6.95 | 28.67 | 6.40 |
| CAZ\|Is.betalactam | 1.62 | 4.39 | 11.93 | 4.33 |
| GEN\|Immunosuppression | 3.28 | 32.67 | 428.46 | 25.79 |
| GEN\|CAZ | 6.67 | 25.98 | 126.93 | 22.55 |
| OFX\|GEN | 9.17 | 45.91 | 375.41 | 38.46 |

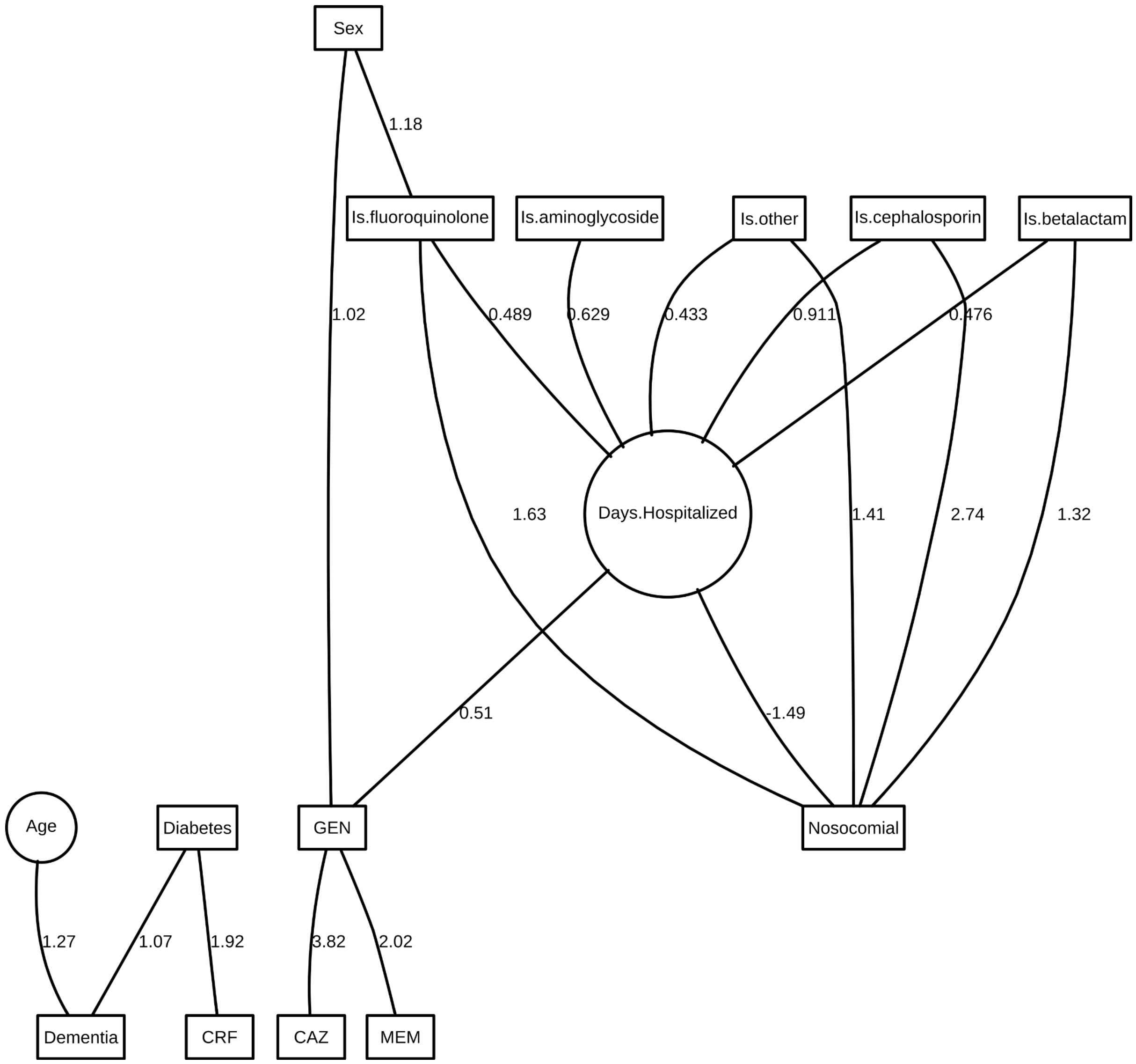

Figure S7: Final DAG for *P aeruginosa* in urine.

Table S9: *P aeruginosa* in urine parameter estimates and their 95% credible intervals. None of the non-intercept credible intervals contains zero.

| | Posterior statistics | | | |
|---|---|---|---|---|
| myvec | 2.50% | median | 97.50% | mode |
| Age\|(Intercept) | -0.07 | 0.00 | 0.07 | 0.00 |
| Age\|precision | 0.90 | 1.00 | 1.10 | 1.00 |
| Sex\|(Intercept) | 0.00 | 0.15 | 0.28 | 0.14 |
| Dementia\|(Intercept) | -3.65 | -3.16 | -2.76 | -3.14 |
| Dementia\|Age | 0.82 | 1.30 | 1.79 | 1.27 |
| Dementia\|Diabetes | 0.46 | 1.08 | 1.62 | 1.07 |
| Diabetes\|(Intercept) | -1.67 | -1.48 | -1.31 | -1.48 |

| | | | | |
|---|---|---|---|---|
| CRF\|(Intercept) | -3.52 | -3.10 | -2.76 | -3.09 |
| CRF\|Diabetes | 1.38 | 1.94 | 2.47 | 1.92 |
| Is.cephalosporin\|(Intercept) | 0.16 | 0.30 | 0.44 | 0.30 |
| Is.betalactam\|(Intercept) | -1.36 | -1.19 | -1.03 | -1.19 |
| Is.aminoglycoside\|(Intercept) | -2.57 | -2.32 | -2.08 | -2.31 |
| Is.fluoroquinolone\|(Intercept) | -4.55 | -3.76 | -3.14 | -3.72 |
| Is.fluoroquinolone\|Sex | 0.44 | 1.21 | 2.01 | 1.18 |
| Is.other\|(Intercept) | -2.35 | -2.11 | -1.90 | -2.11 |
| Days.Hospitalized\|(Intercept) | -0.85 | -0.76 | -0.68 | -0.76 |
| Days.Hospitalized\|Is.cephalosporin | 0.80 | 0.91 | 1.02 | 0.91 |
| Days.Hospitalized\|Is.betalactam | 0.35 | 0.47 | 0.60 | 0.48 |
| Days.Hospitalized\|Is.aminoglycoside | 0.43 | 0.62 | 0.81 | 0.63 |
| Days.Hospitalized\|Is.fluoroquinolone | 0.23 | 0.48 | 0.73 | 0.49 |
| Days.Hospitalized\|Is.other | 0.25 | 0.43 | 0.61 | 0.43 |
| Days.Hospitalized\|precision | 1.61 | 1.77 | 1.95 | 1.79 |
| Nosocomial\|(Intercept) | -2.83 | -2.45 | -2.10 | -2.43 |
| Nosocomial\|Is.cephalosporin | 2.30 | 2.76 | 3.25 | 2.74 |
| Nosocomial\|Is.betalactam | 0.89 | 1.32 | 1.77 | 1.32 |
| Nosocomial\|Is.fluoroquinolone | 0.76 | 1.66 | 2.53 | 1.63 |
| Nosocomial\|Is.other | 0.82 | 1.43 | 2.02 | 1.41 |
| Nosocomial\|Days.Hospitalized | -1.79 | -1.50 | -1.24 | -1.49 |
| CAZ\|(Intercept) | -5.03 | -4.31 | -3.71 | -4.26 |
| CAZ\|GEN | 3.10 | 3.84 | 4.64 | 3.82 |
| GEN\|(Intercept) | -3.19 | -2.74 | -2.35 | -2.72 |
| GEN\|Sex | 0.55 | 1.02 | 1.53 | 1.02 |
| GEN\|Days.Hospitalized | 0.30 | 0.51 | 0.72 | 0.51 |
| MEM\|(Intercept) | -4.56 | -3.94 | -3.44 | -3.92 |
| MEM\|GEN | 1.19 | 2.04 | 2.82 | 2.02 |
| | OR | | | |
| | 2.50% | median | 97.50% | mode |
| Dementia\|Age | 2.27 | 3.66 | 6.02 | 3.57 |
| Dementia\|Diabetes | 1.59 | 2.94 | 5.08 | 2.91 |
| CRF\|Diabetes | 3.97 | 6.95 | 11.88 | 6.85 |
| Is.fluoroquinolone\|Sex | 1.55 | 3.35 | 7.46 | 3.24 |
| Nosocomial\|Is.cephalosporin | 9.96 | 15.87 | 25.77 | 15.55 |
| Nosocomial\|Is.betalactam | 2.44 | 3.75 | 5.88 | 3.73 |

| | | | | |
|---|---|---|---|---|
| Nosocomial\|Is.fluoroquinolone | 2.14 | 5.27 | 12.54 | 5.12 |
| Nosocomial\|Is.other | 2.28 | 4.20 | 7.55 | 4.10 |
| Nosocomial\|Days.Hospitalized | 0.17 | 0.22 | 0.29 | 0.23 |
| CAZ\|GEN | 22.13 | 46.39 | 103.41 | 45.41 |
| GEN\|Sex | 1.74 | 2.78 | 4.62 | 2.77 |
| GEN\|Days.Hospitalized | 1.36 | 1.67 | 2.05 | 1.67 |
| MEM\|GEN | 3.28 | 7.67 | 16.78 | 7.54 |

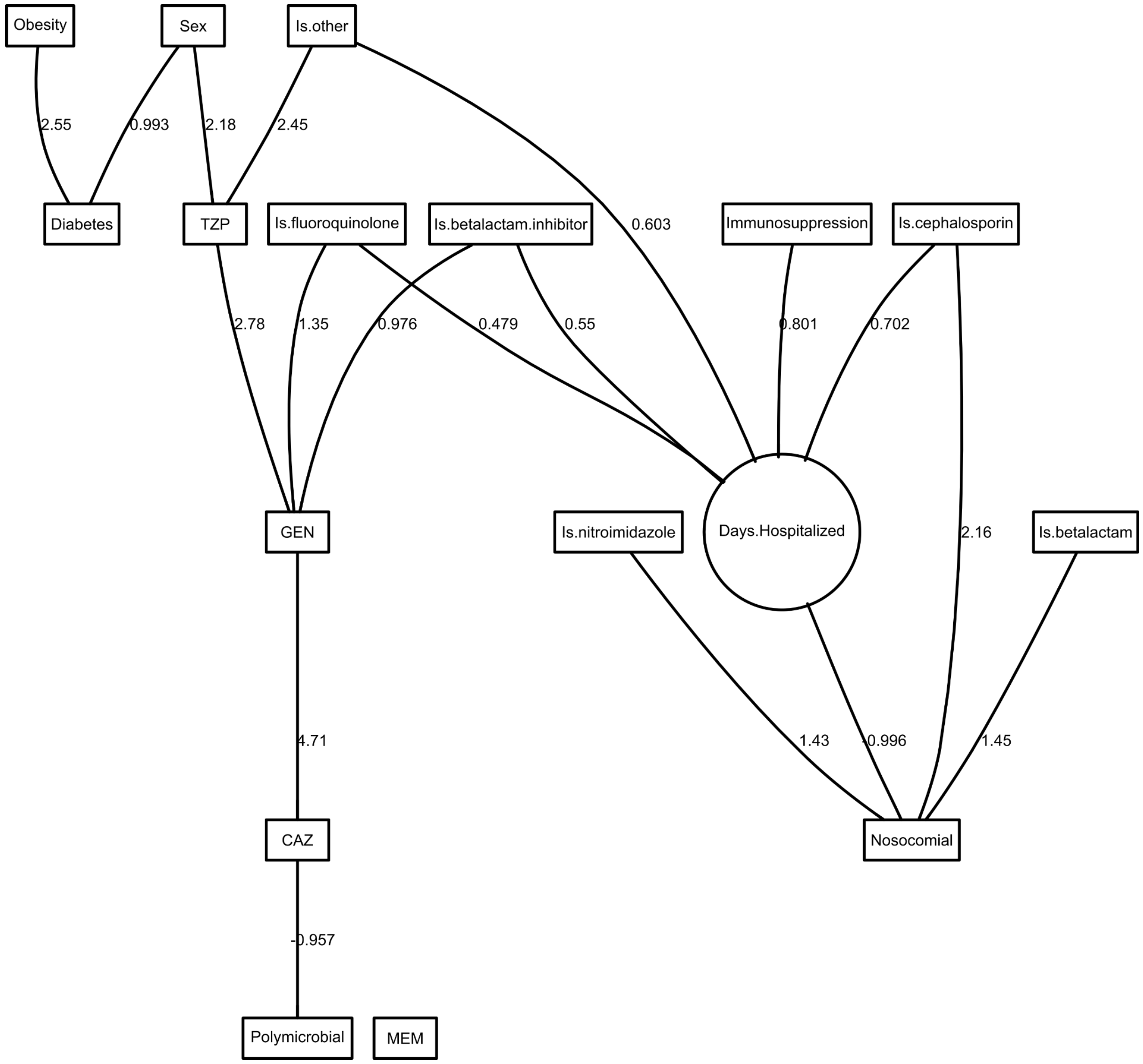

Figure S8: Final DAG for *P aeruginosa* in wound.

Table S10: *P aeruginosa* in wound parameter estimates and their 95% credible intervals. None of the non-intercept credible intervals contains zero.

| | Posterior statistics | | | |
|---|---|---|---|---|
| myvec | 2.50% | median | 97.50% | mode |
| Sex\|(Intercept) | 0.04 | 0.22 | 0.38 | 0.21 |
| Immunosuppression\|(Intercept) | -4.02 | -3.47 | -3.01 | -3.45 |
| Diabetes\|(Intercept) | -2.34 | -1.92 | -1.56 | -1.91 |
| Diabetes\|Sex | 0.54 | 0.99 | 1.45 | 0.99 |
| Diabetes\|Obesity | 1.71 | 2.58 | 3.52 | 2.55 |
| Obesity\|(Intercept) | -3.28 | -2.88 | -2.53 | -2.86 |
| Is.cephalosporin\|(Intercept) | 0.48 | 0.66 | 0.84 | 0.66 |
| Is.betalactam\|(Intercept) | -0.96 | -0.77 | -0.60 | -0.78 |
| Is.betalactam.inhibitor\|(Intercept) | -1.27 | -1.07 | -0.88 | -1.07 |
| Is.nitroimidazole\|(Intercept) | -1.71 | -1.48 | -1.27 | -1.48 |
| Is.fluoroquinolone\|(Intercept) | -2.32 | -2.04 | -1.79 | -2.04 |
| Is.other\|(Intercept) | -1.96 | -1.72 | -1.49 | -1.71 |
| Days.Hospitalized\|(Intercept) | -0.90 | -0.77 | -0.65 | -0.78 |
| Days.Hospitalized\|Immunosuppression | 0.39 | 0.79 | 1.20 | 0.80 |
| Days.Hospitalized\|Is.cephalosporin | 0.55 | 0.70 | 0.85 | 0.70 |
| Days.Hospitalized\|Is.betalactam.inhibitor | 0.39 | 0.55 | 0.71 | 0.55 |
| Days.Hospitalized\|Is.fluoroquinolone | 0.25 | 0.47 | 0.70 | 0.48 |
| Days.Hospitalized\|Is.other | 0.39 | 0.60 | 0.80 | 0.60 |
| Days.Hospitalized\|precision | 1.33 | 1.52 | 1.70 | 1.53 |
| Nosocomial\|(Intercept) | -1.49 | -1.07 | -0.69 | -1.06 |
| Nosocomial\|Is.cephalosporin | 1.63 | 2.18 | 2.74 | 2.16 |
| Nosocomial\|Is.betalactam | 0.95 | 1.48 | 2.01 | 1.45 |
| Nosocomial\|Is.nitroimidazole | 0.72 | 1.48 | 2.27 | 1.43 |
| Nosocomial\|Days.Hospitalized | -1.31 | -1.00 | -0.74 | -1.00 |
| Polymicrobial\|(Intercept) | 0.48 | 0.67 | 0.86 | 0.67 |
| Polymicrobial\|CAZ | -1.55 | -0.97 | -0.41 | -0.96 |
| CAZ\|(Intercept) | -5.68 | -4.58 | -3.78 | -4.51 |
| CAZ\|GEN | 3.84 | 4.79 | 5.92 | 4.71 |
| GEN\|(Intercept) | -2.70 | -2.32 | -2.00 | -2.32 |
| GEN\|Is.betalactam.inhibitor | 0.44 | 0.99 | 1.49 | 0.98 |
| GEN\|Is.fluoroquinolone | 0.70 | 1.36 | 1.98 | 1.35 |
| GEN\|TZP | 1.78 | 2.86 | 4.03 | 2.78 |
| MEM\|(Intercept) | -3.80 | -3.30 | -2.88 | -3.29 |
| TZP\|(Intercept) | -8.01 | -5.95 | -4.56 | -5.71 |

| | | | | |
|---|---|---|---|---|
| TZP\|Sex | 0.92 | 2.31 | 4.28 | 2.18 |
| TZP\|Is.other | 1.52 | 2.48 | 3.45 | 2.45 |
| | OR | | | |
| | 2.50% | median | 97.50% | mode |
| Diabetes\|Sex | 1.71 | 2.70 | 4.27 | 2.70 |
| Diabetes\|Obesity | 5.54 | 13.20 | 33.69 | 12.75 |
| Nosocomial\|Is.cephalosporin | 5.12 | 8.88 | 15.41 | 8.71 |
| Nosocomial\|Is.betalactam | 2.58 | 4.38 | 7.46 | 4.27 |
| Nosocomial\|Is.nitroimidazole | 2.06 | 4.40 | 9.72 | 4.20 |
| Nosocomial\|Days.Hospitalized | 0.27 | 0.37 | 0.48 | 0.37 |
| Polymicrobial\|CAZ | 0.21 | 0.38 | 0.66 | 0.38 |
| CAZ\|GEN | 46.73 | 119.80 | 373.68 | 110.76 |
| GEN\|Is.betalactam.inhibitor | 1.56 | 2.68 | 4.42 | 2.65 |
| GEN\|Is.fluoroquinolone | 2.01 | 3.91 | 7.22 | 3.86 |
| GEN\|TZP | 5.92 | 17.41 | 56.00 | 16.13 |
| TZP\|Sex | 2.52 | 10.04 | 72.55 | 8.80 |
| TZP\|Is.other | 4.56 | 11.99 | 31.52 | 11.64 |

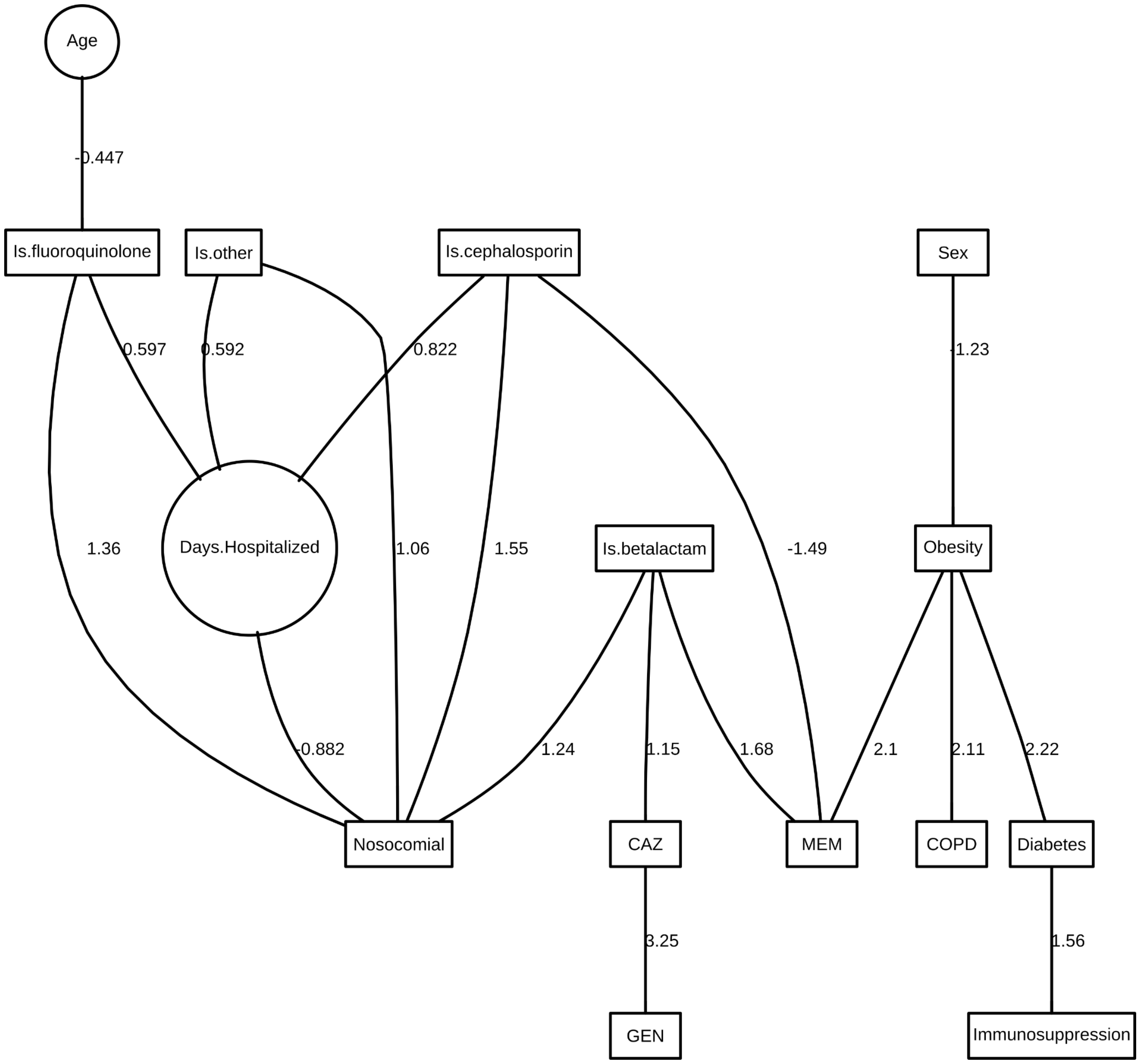

Figure S9: Final DAG for *P aeruginosa* in sputum.

Table S11: *P aeruginosa* in sputum parameter estimates and their 95% credible intervals. None of the non-intercept credible intervals contains zero.

| | Posterior statistics | | | |
|---|---|---|---|---|
| myvec | 2.50% | median | 97.50% | mode |
| Age\|(Intercept) | -0.10 | 0.00 | 0.09 | 0.00 |
| Age\|precision | 0.87 | 1.00 | 1.14 | 1.00 |
| Sex\|(Intercept) | 0.42 | 0.62 | 0.82 | 0.62 |
| Immunosuppression\|(Intercept) | -4.11 | -3.46 | -2.90 | -3.42 |
| Immunosuppression\|Diabetes | 0.63 | 1.57 | 2.44 | 1.56 |
| Diabetes\|(Intercept) | -2.15 | -1.85 | -1.58 | -1.84 |
| Diabetes\|Obesity | 1.41 | 2.25 | 3.09 | 2.22 |

| | | | | |
|---|---|---|---|---|
| COPD\|(Intercept) | -2.37 | -2.04 | -1.76 | -2.04 |
| COPD\|Obesity | 1.26 | 2.11 | 2.94 | 2.11 |
| Obesity\|(Intercept) | -2.70 | -2.13 | -1.65 | -2.12 |
| Obesity\|Sex | -2.11 | -1.24 | -0.44 | -1.23 |
| Is.cephalosporin\|(Intercept) | 0.33 | 0.53 | 0.71 | 0.53 |
| Is.betalactam\|(Intercept) | -0.91 | -0.70 | -0.51 | -0.70 |
| Is.fluoroquinolone\|(Intercept) | -2.12 | -1.82 | -1.56 | -1.82 |
| Is.fluoroquinolone\|Age | -0.69 | -0.45 | -0.21 | -0.45 |
| Is.other\|(Intercept) | -1.14 | -0.92 | -0.72 | -0.92 |
| Days.Hospitalized\|(Intercept) | -0.91 | -0.77 | -0.65 | -0.77 |
| Days.Hospitalized\|Is.cephalosporin | 0.66 | 0.82 | 0.98 | 0.82 |
| Days.Hospitalized\|Is.fluoroquinolone | 0.38 | 0.59 | 0.81 | 0.60 |
| Days.Hospitalized\|Is.other | 0.42 | 0.59 | 0.76 | 0.59 |
| Days.Hospitalized\|precision | 1.32 | 1.51 | 1.73 | 1.53 |
| Nosocomial\|(Intercept) | -1.18 | -0.71 | -0.29 | -0.71 |
| Nosocomial\|Is.cephalosporin | 1.01 | 1.57 | 2.16 | 1.55 |
| Nosocomial\|Is.betalactam | 0.70 | 1.27 | 1.84 | 1.24 |
| Nosocomial\|Is.fluoroquinolone | 0.59 | 1.42 | 2.27 | 1.36 |
| Nosocomial\|Is.other | 0.49 | 1.08 | 1.72 | 1.06 |
| Nosocomial\|Days.Hospitalized | -1.23 | -0.89 | -0.60 | -0.88 |
| CAZ\|(Intercept) | -3.67 | -3.05 | -2.54 | -3.02 |
| CAZ\|Is.betalactam | 0.41 | 1.17 | 1.89 | 1.15 |
| GEN\|(Intercept) | -4.34 | -3.65 | -3.09 | -3.61 |
| GEN\|CAZ | 2.33 | 3.28 | 4.22 | 3.25 |
| MEM\|(Intercept) | -4.26 | -3.31 | -2.56 | -3.23 |
| MEM\|Obesity | 0.75 | 2.12 | 3.34 | 2.10 |
| MEM\|Is.cephalosporin | -2.55 | -1.54 | -0.60 | -1.49 |
| MEM\|Is.betalactam | 0.73 | 1.70 | 2.76 | 1.68 |
| | OR | | | |
| | 2.50% | median | 97.50% | mode |
| Immunosuppression\|Diabetes | 1.88 | 4.82 | 11.52 | 4.76 |
| Diabetes\|Obesity | 4.08 | 9.45 | 21.90 | 9.20 |
| COPD\|Obesity | 3.51 | 8.29 | 18.94 | 8.26 |
| Obesity\|Sex | 0.12 | 0.29 | 0.65 | 0.29 |
| Is.fluoroquinolone\|Age | 0.50 | 0.64 | 0.81 | 0.64 |
| Nosocomial\|Is.cephalosporin | 2.74 | 4.81 | 8.66 | 4.72 |

| | | | | |
|---|---|---|---|---|
| Nosocomial\|Is.betalactam | 2.01 | 3.56 | 6.30 | 3.46 |
| Nosocomial\|Is.fluoroquinolone | 1.80 | 4.12 | 9.73 | 3.91 |
| Nosocomial\|Is.other | 1.62 | 2.93 | 5.56 | 2.89 |
| Nosocomial\|Days.Hospitalized | 0.29 | 0.41 | 0.55 | 0.41 |
| CAZ\|Is.betalactam | 1.51 | 3.21 | 6.64 | 3.16 |
| GEN\|CAZ | 10.31 | 26.52 | 68.21 | 25.87 |
| MEM\|Obesity | 2.12 | 8.30 | 28.09 | 8.16 |
| MEM\|Is.cephalosporin | 0.08 | 0.21 | 0.55 | 0.23 |
| MEM\|Is.betalactam | 2.07 | 5.48 | 15.77 | 5.34 |

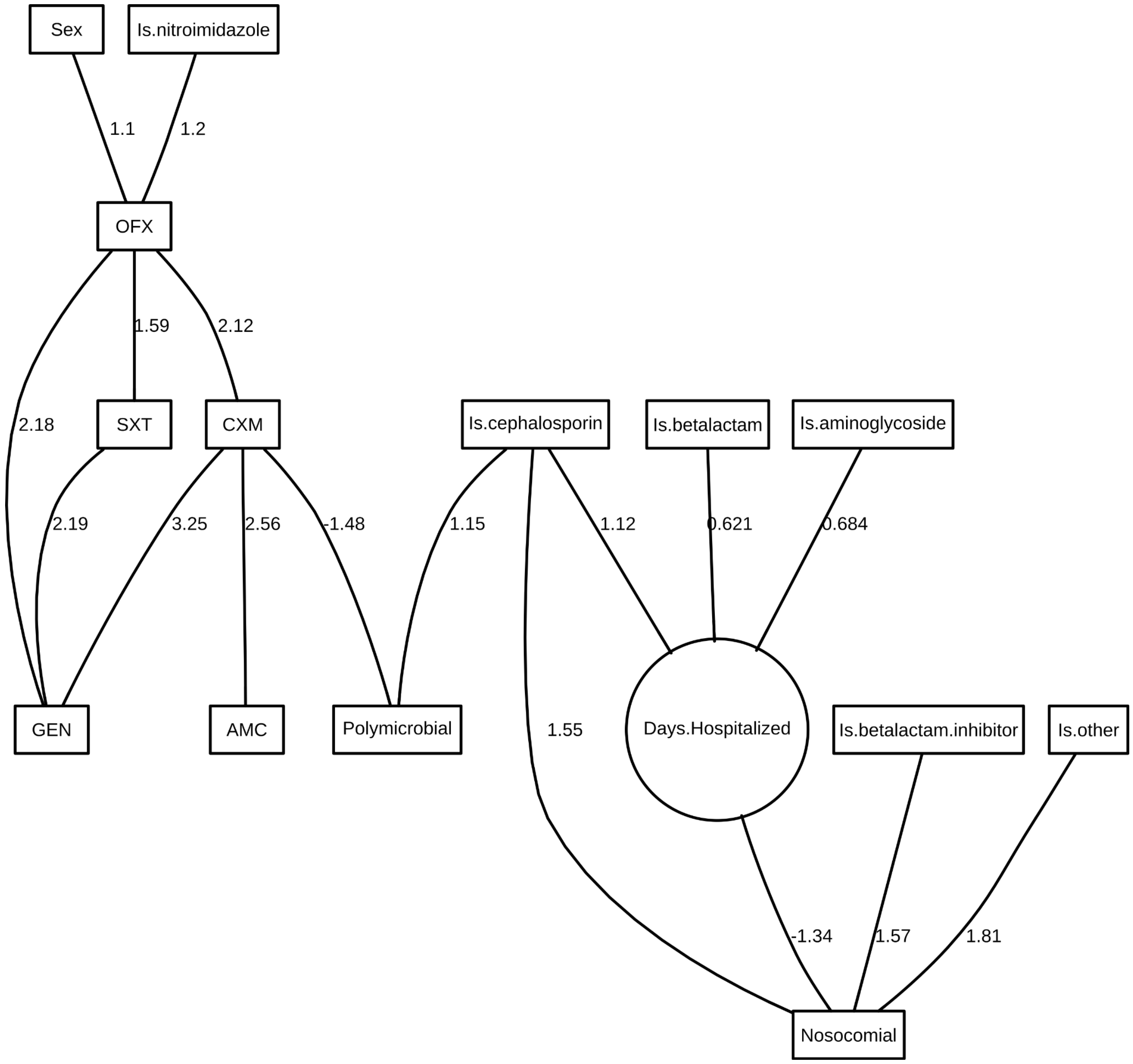

Figure S10: Final DAG for *P mirabilis* in urine.

Table S12: *P mirabilis* in urine parameter estimates and their 95% credible intervals. None of the non-intercept credible intervals contains zero.

| | Posterior statistics | | | |
|---|---|---|---|---|
| myvec | 2.50% | median | 97.50% | mode |
| Sex\|(Intercept) | -0.37 | -0.15 | 0.07 | -0.14 |
| Is.cephalosporin\|(Intercept) | -0.77 | -0.53 | -0.31 | -0.53 |
| Is.betalactam\|(Intercept) | -1.86 | -1.55 | -1.29 | -1.55 |
| Is.betalactam.inhibitor\|(Intercept) | -2.19 | -1.85 | -1.55 | -1.85 |
| Is.nitroimidazole\|(Intercept) | -2.65 | -2.25 | -1.90 | -2.24 |
| Is.aminoglycoside\|(Intercept) | -2.65 | -2.25 | -1.90 | -2.24 |
| Is.other\|(Intercept) | -2.73 | -2.32 | -1.96 | -2.31 |
| Days.Hospitalized\|(Intercept) | -0.69 | -0.59 | -0.49 | -0.59 |
| Days.Hospitalized\|Is.cephalosporin | 0.94 | 1.11 | 1.28 | 1.12 |
| Days.Hospitalized\|Is.betalactam | 0.41 | 0.62 | 0.83 | 0.62 |
| Days.Hospitalized\|Is.aminoglycoside | 0.41 | 0.68 | 0.94 | 0.68 |
| Days.Hospitalized\|precision | 1.83 | 2.16 | 2.50 | 2.18 |
| Nosocomial\|(Intercept) | -2.55 | -2.03 | -1.55 | -1.98 |
| Nosocomial\|Is.cephalosporin | 0.81 | 1.60 | 2.38 | 1.55 |
| Nosocomial\|Is.betalactam.inhibitor | 0.68 | 1.59 | 2.54 | 1.57 |
| Nosocomial\|Is.other | 0.85 | 1.86 | 2.90 | 1.81 |
| Nosocomial\|Days.Hospitalized | -1.94 | -1.37 | -0.90 | -1.34 |
| Polymicrobial\|(Intercept) | -2.27 | -1.80 | -1.40 | -1.79 |
| Polymicrobial\|Is.cephalosporin | 0.54 | 1.17 | 1.76 | 1.15 |
| Polymicrobial\|CXM | -2.61 | -1.53 | -0.67 | -1.48 |
| AMC\|(Intercept) | -4.27 | -3.43 | -2.77 | -3.39 |
| AMC\|CXM | 1.75 | 2.59 | 3.50 | 2.56 |
| CXM\|(Intercept) | -2.75 | -2.23 | -1.77 | -2.21 |
| CXM\|OFX | 1.56 | 2.15 | 2.74 | 2.12 |
| GEN\|(Intercept) | -6.98 | -5.35 | -4.14 | -5.20 |
| GEN\|CXM | 2.48 | 3.34 | 4.36 | 3.25 |
| GEN\|OFX | 1.31 | 2.26 | 3.29 | 2.18 |
| GEN\|SXT | 1.37 | 2.28 | 3.26 | 2.19 |
| OFX\|(Intercept) | -1.32 | -0.97 | -0.64 | -0.96 |
| OFX\|Sex | 0.63 | 1.12 | 1.59 | 1.10 |
| OFX\|Is.nitroimidazole | 0.39 | 1.23 | 2.11 | 1.20 |
| SXT\|(Intercept) | -1.65 | -1.28 | -0.94 | -1.27 |

| | | | | |
|---|---|---|---|---|
| SXT\|OFX | 1.10 | 1.60 | 2.08 | 1.59 |
| | OR | | | |
| | 2.50% | median | 97.50% | mode |
| Nosocomial\|Is.cephalosporin | 2.26 | 4.95 | 10.85 | 4.73 |
| Nosocomial\|Is.betalactam.inhibitor | 1.98 | 4.93 | 12.72 | 4.81 |
| Nosocomial\|Is.other | 2.33 | 6.39 | 18.24 | 6.14 |
| Nosocomial\|Days.Hospitalized | 0.14 | 0.25 | 0.41 | 0.26 |
| Polymicrobial\|Is.cephalosporin | 1.71 | 3.23 | 5.81 | 3.17 |
| Polymicrobial\|CXM | 0.07 | 0.22 | 0.51 | 0.23 |
| AMC\|CXM | 5.78 | 13.34 | 33.00 | 12.88 |
| CXM\|OFX | 4.74 | 8.55 | 15.43 | 8.35 |
| GEN\|CXM | 11.90 | 28.33 | 77.97 | 25.88 |
| GEN\|OFX | 3.70 | 9.57 | 26.79 | 8.84 |
| GEN\|SXT | 3.95 | 9.75 | 25.94 | 8.96 |
| OFX\|Sex | 1.88 | 3.06 | 4.89 | 3.01 |
| OFX\|Is.nitroimidazole | 1.47 | 3.43 | 8.26 | 3.31 |
| SXT\|OFX | 3.00 | 4.95 | 8.01 | 4.90 |

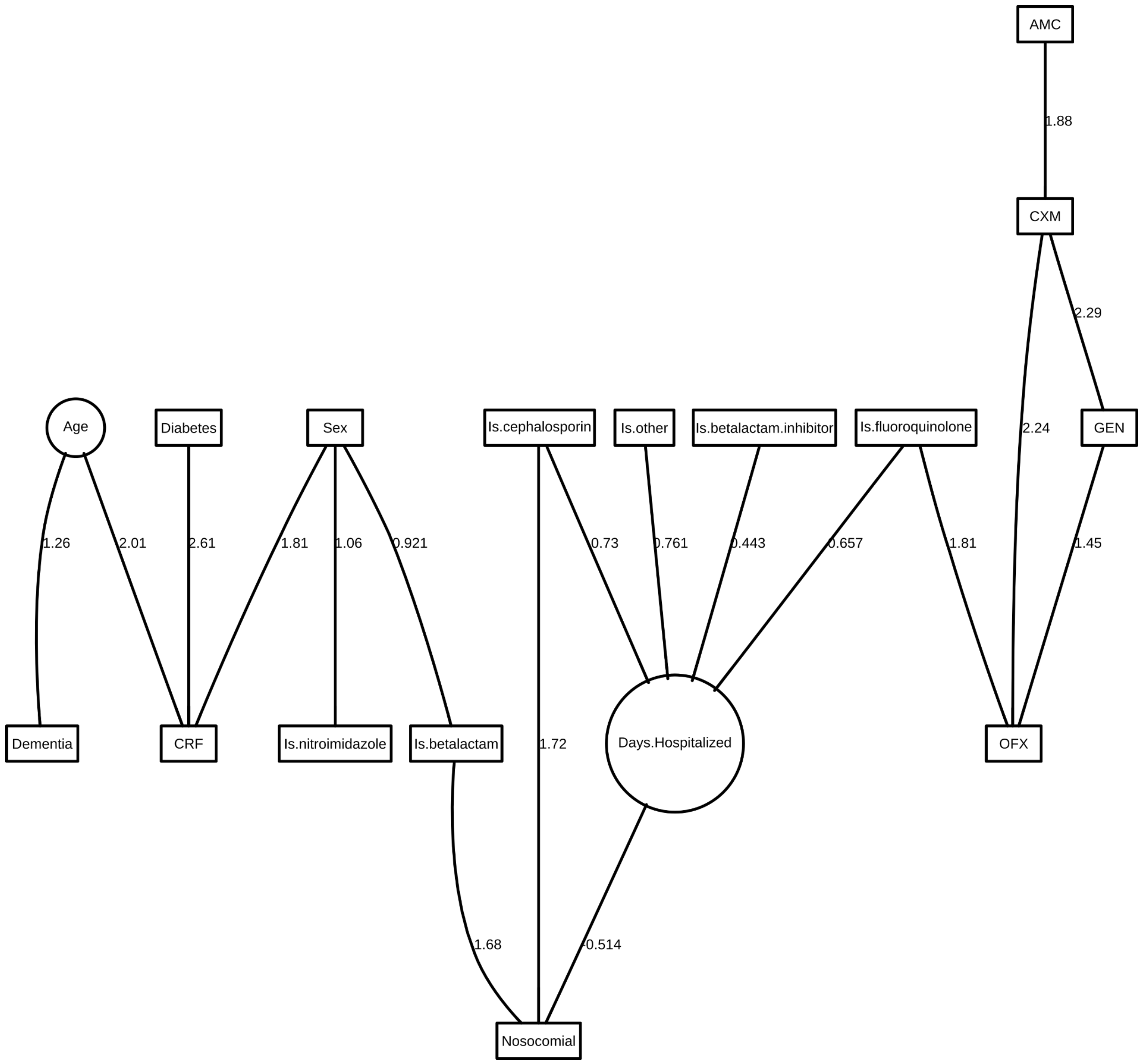

Figure S11: Final DAG for *P mirabilis* in wound.

Table S13: *P mirabilis* in wound parameter estimates and their 95% credible intervals. None of the non-intercept credible intervals contains zero.

| | Posterior statistics | | | |
|---|---|---|---|---|
| myvec | 2.50% | median | 97.50% | mode |
| Age\|(Intercept) | -0.12 | 0.00 | 0.11 | 0.00 |
| Age\|precision | 0.85 | 1.00 | 1.17 | 1.00 |
| Sex\|(Intercept) | -0.21 | 0.02 | 0.24 | 0.02 |
| Dementia\|(Intercept) | -5.18 | -4.00 | -3.17 | -3.90 |
| Dementia\|Age | 0.46 | 1.31 | 2.27 | 1.26 |
| Diabetes\|(Intercept) | -1.28 | -1.01 | -0.77 | -1.01 |
| CRF\|(Intercept) | -7.41 | -5.62 | -4.30 | -5.43 |

| | | | | |
|---|---|---|---|---|
| CRF\|Age | 1.26 | 2.09 | 3.02 | 2.01 |
| CRF\|Sex | 0.84 | 1.89 | 3.12 | 1.81 |
| CRF\|Diabetes | 1.67 | 2.72 | 3.82 | 2.61 |
| Is.cephalosporin\|(Intercept) | -0.02 | 0.20 | 0.43 | 0.21 |
| Is.betalactam\|(Intercept) | -1.23 | -0.86 | -0.53 | -0.86 |
| Is.betalactam\|Sex | 0.45 | 0.92 | 1.39 | 0.92 |
| Is.betalactam.inhibitor\|(Intercept) | -0.72 | -0.48 | -0.25 | -0.48 |
| Is.nitroimidazole\|(Intercept) | -3.09 | -2.44 | -1.91 | -2.41 |
| Is.nitroimidazole\|Sex | 0.37 | 1.06 | 1.79 | 1.06 |
| Is.fluoroquinolone\|(Intercept) | -2.48 | -2.11 | -1.77 | -2.09 |
| Is.other\|(Intercept) | -2.19 | -1.84 | -1.54 | -1.83 |
| Days.Hospitalized\|(Intercept) | -0.89 | -0.75 | -0.61 | -0.75 |
| Days.Hospitalized\|Is.cephalosporin | 0.55 | 0.73 | 0.90 | 0.73 |
| Days.Hospitalized\|Is.betalactam.inhibitor | 0.26 | 0.44 | 0.62 | 0.44 |
| Days.Hospitalized\|Is.fluoroquinolone | 0.35 | 0.65 | 0.95 | 0.66 |
| Days.Hospitalized\|Is.other | 0.49 | 0.76 | 1.02 | 0.76 |
| Days.Hospitalized\|precision | 1.44 | 1.70 | 1.98 | 1.72 |
| Nosocomial\|(Intercept) | -1.70 | -1.19 | -0.74 | -1.18 |
| Nosocomial\|Is.cephalosporin | 1.14 | 1.74 | 2.37 | 1.72 |
| Nosocomial\|Is.betalactam | 1.11 | 1.70 | 2.29 | 1.68 |
| Nosocomial\|Days.Hospitalized | -0.87 | -0.52 | -0.21 | -0.51 |
| AMC\|(Intercept) | -2.52 | -2.14 | -1.80 | -2.13 |
| CXM\|(Intercept) | -2.00 | -1.66 | -1.35 | -1.65 |
| CXM\|AMC | 1.11 | 1.90 | 2.66 | 1.88 |
| GEN\|(Intercept) | -2.74 | -2.26 | -1.88 | -2.26 |
| GEN\|CXM | 1.64 | 2.31 | 2.97 | 2.29 |
| OFX\|(Intercept) | -2.84 | -2.35 | -1.92 | -2.32 |
| OFX\|Is.fluoroquinolone | 0.91 | 1.83 | 2.70 | 1.81 |
| OFX\|CXM | 1.55 | 2.29 | 2.99 | 2.24 |
| OFX\|GEN | 0.68 | 1.48 | 2.24 | 1.45 |
| | OR | | | |
| | 2.50% | median | 97.50% | mode |
| Dementia\|Age | 1.58 | 3.71 | 9.68 | 3.51 |
| CRF\|Age | 3.52 | 8.08 | 20.52 | 7.48 |
| CRF\|Sex | 2.31 | 6.62 | 22.67 | 6.13 |
| CRF\|Diabetes | 5.30 | 15.21 | 45.54 | 13.58 |

| | | | | |
|---|---|---|---|---|
| Is.betalactam\|Sex | 1.57 | 2.51 | 4.01 | 2.51 |
| Is.nitroimidazole\|Sex | 1.45 | 2.90 | 5.96 | 2.87 |
| Nosocomial\|Is.cephalosporin | 3.11 | 5.71 | 10.74 | 5.57 |
| Nosocomial\|Is.betalactam | 3.03 | 5.48 | 9.91 | 5.35 |
| Nosocomial\|Days.Hospitalized | 0.42 | 0.60 | 0.81 | 0.60 |
| CXM\|AMC | 3.03 | 6.68 | 14.30 | 6.58 |
| GEN\|CXM | 5.17 | 10.05 | 19.55 | 9.88 |
| OFX\|Is.fluoroquinolone | 2.49 | 6.21 | 14.92 | 6.10 |
| OFX\|CXM | 4.71 | 9.84 | 19.97 | 9.43 |
| OFX\|GEN | 1.97 | 4.37 | 9.43 | 4.26 |

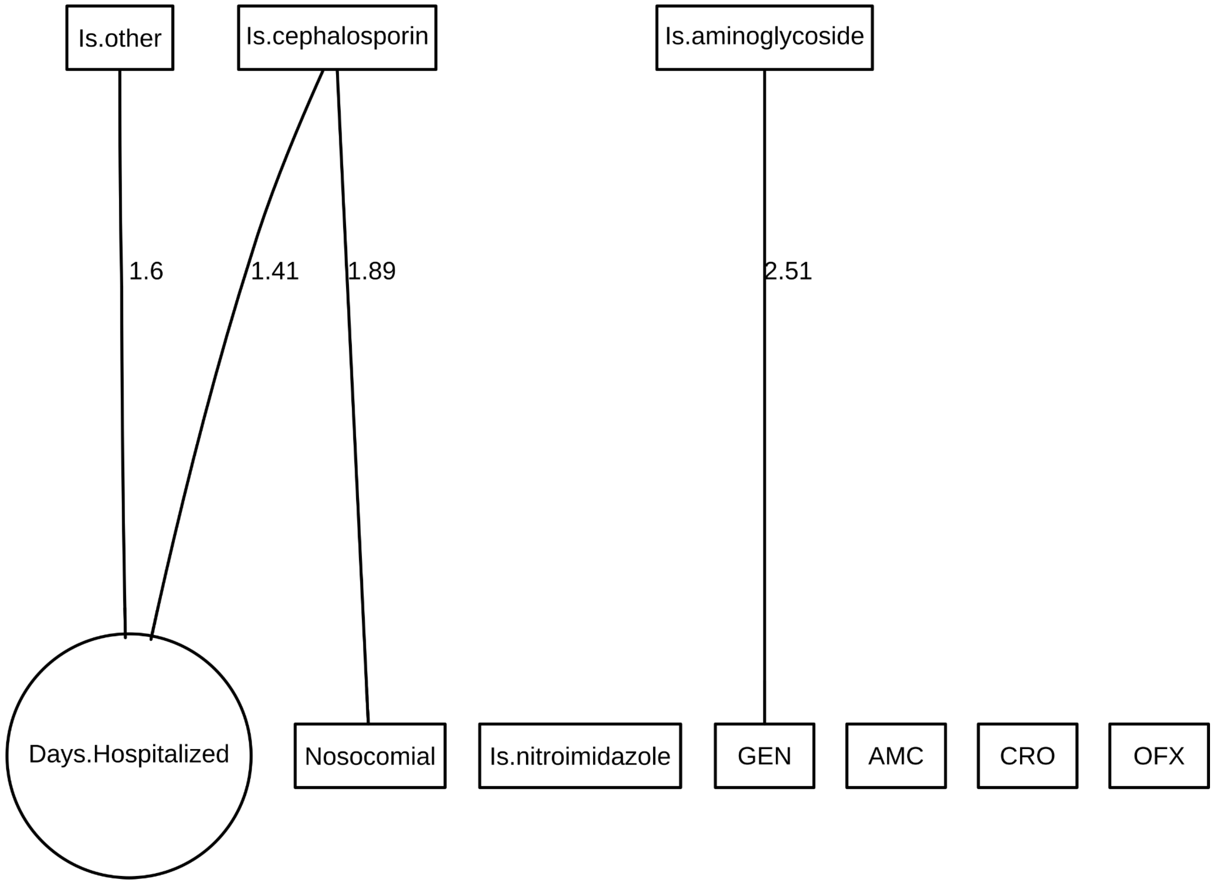

Figure S12: Final DAG for *P mirabilis* in aerobic blood.

Table S14: *P mirabilis* in aerobic blood parameter estimates and their 95% credible intervals. None of the non-intercept credible intervals contains zero.

| | Posterior statistics | | | |
|---|---|---|---|---|
| myvec | 2.50% | median | 97.50% | mode |
| Is.cephalosporin\|(Intercept) | -0.84 | -0.25 | 0.31 | -0.24 |
| Is.nitroimidazole\|(Intercept) | -3.06 | -2.04 | -1.27 | -1.99 |
| Is.aminoglycoside\|(Intercept) | -3.38 | -2.25 | -1.40 | -2.20 |
| Is.other\|(Intercept) | -4.36 | -2.86 | -1.81 | -2.75 |
| Days.Hospitalized\|(Intercept) | -0.97 | -0.71 | -0.47 | -0.72 |
| Days.Hospitalized\|Is.cephalosporin | 1.04 | 1.40 | 1.76 | 1.41 |
| Days.Hospitalized\|Is.other | 0.82 | 1.59 | 2.36 | 1.60 |
| Days.Hospitalized\|precision | 1.64 | 2.59 | 3.74 | 2.67 |
| Nosocomial\|(Intercept) | -2.72 | -1.59 | -0.68 | -1.53 |
| Nosocomial\|Is.cephalosporin | 0.65 | 2.00 | 3.39 | 1.89 |
| AMC\|(Intercept) | -2.80 | -1.84 | -1.11 | -1.81 |
| CRO\|(Intercept) | -1.77 | -1.06 | -0.45 | -1.05 |
| GEN\|(Intercept) | -1.91 | -1.14 | -0.49 | -1.13 |
| GEN\|Is.aminoglycoside | 0.52 | 2.81 | 6.19 | 2.51 |
| OFX\|(Intercept) | -2.38 | -1.54 | -0.85 | -1.52 |
| | OR | | | |
| | 2.50% | median | 97.50% | mode |
| Nosocomial\|Is.cephalosporin | 1.92 | 7.36 | 29.65 | 6.64 |
| GEN\|Is.aminoglycoside | 1.69 | 16.66 | 489.93 | 12.32 |

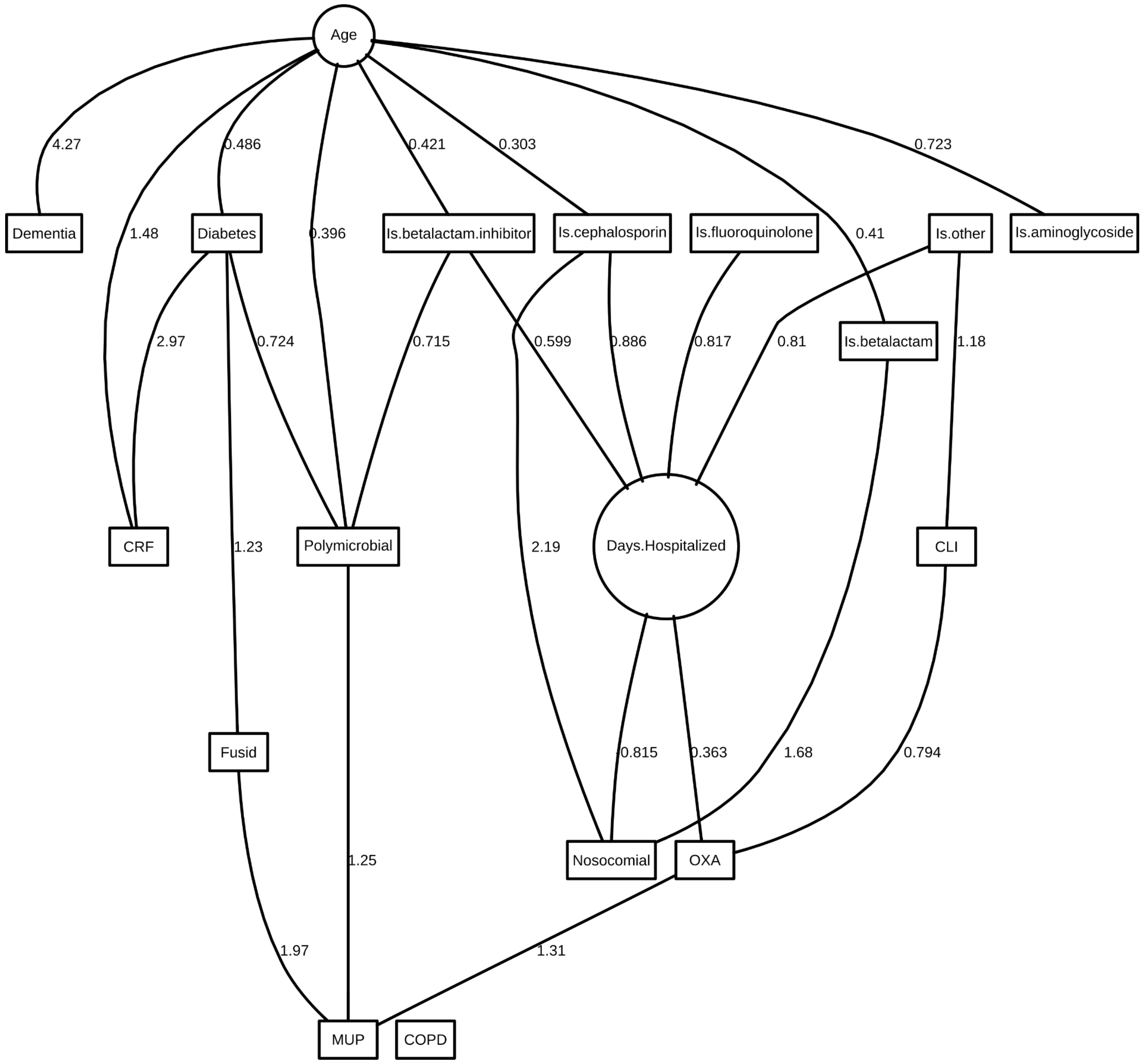

Figure S13: Final DAG for *S aureus* in wound.

Table S15: *S aureus* in wound parameter estimates and their 95% credible intervals. None of the non-intercept credible intervals contains zero.

| | Posterior statistics | | | |
|---|---|---|---|---|
| myvec | 2.50% | median | 97.50% | mode |
| Age\|(Intercept) | -0.08 | 0.00 | 0.07 | 0.00 |
| Age\|precision | 0.90 | 1.00 | 1.11 | 1.00 |
| Dementia\|(Intercept) | -13.95 | -9.26 | -6.31 | -8.86 |
| Dementia\|Age | 2.28 | 4.55 | 7.59 | 4.27 |
| Diabetes\|(Intercept) | -1.83 | -1.62 | -1.42 | -1.61 |
| Diabetes\|Age | 0.27 | 0.49 | 0.71 | 0.49 |
| COPD\|(Intercept) | -4.18 | -3.66 | -3.24 | -3.65 |

| | | | | |
|---|---|---|---|---|
| CRF\|(Intercept) | -6.30 | -5.18 | -4.33 | -5.09 |
| CRF\|Age | 0.84 | 1.52 | 2.26 | 1.48 |
| CRF\|Diabetes | 2.17 | 3.03 | 3.96 | 2.97 |
| Is.cephalosporin\|(Intercept) | -0.46 | -0.31 | -0.16 | -0.31 |
| Is.cephalosporin\|Age | 0.14 | 0.31 | 0.46 | 0.30 |
| Is.betalactam\|(Intercept) | -1.34 | -1.16 | -0.99 | -1.15 |
| Is.betalactam\|Age | 0.22 | 0.42 | 0.60 | 0.41 |
| Is.betalactam.inhibitor\|(Intercept) | -1.40 | -1.22 | -1.05 | -1.22 |
| Is.betalactam.inhibitor\|Age | 0.23 | 0.42 | 0.62 | 0.42 |
| Is.aminoglycoside\|(Intercept) | -3.98 | -3.45 | -3.03 | -3.42 |
| Is.aminoglycoside\|Age | 0.27 | 0.74 | 1.24 | 0.72 |
| Is.fluoroquinolone\|(Intercept) | -2.89 | -2.58 | -2.32 | -2.58 |
| Is.other\|(Intercept) | -2.51 | -2.25 | -2.01 | -2.24 |
| Days.Hospitalized\|(Intercept) | -0.73 | -0.65 | -0.58 | -0.65 |
| Days.Hospitalized\|Is.cephalosporin | 0.78 | 0.88 | 0.99 | 0.89 |
| Days.Hospitalized\|Is.betalactam.inhibitor | 0.48 | 0.60 | 0.72 | 0.60 |
| Days.Hospitalized\|Is.fluoroquinolone | 0.60 | 0.81 | 1.02 | 0.82 |
| Days.Hospitalized\|Is.other | 0.63 | 0.81 | 0.98 | 0.81 |
| Days.Hospitalized\|precision | 1.88 | 2.09 | 2.31 | 2.10 |
| Nosocomial\|(Intercept) | -1.83 | -1.54 | -1.27 | -1.53 |
| Nosocomial\|Is.cephalosporin | 1.76 | 2.20 | 2.65 | 2.19 |
| Nosocomial\|Is.betalactam | 1.25 | 1.69 | 2.12 | 1.68 |
| Nosocomial\|Days.Hospitalized | -1.07 | -0.82 | -0.59 | -0.82 |
| Polymicrobial\|(Intercept) | -0.90 | -0.71 | -0.52 | -0.70 |
| Polymicrobial\|Age | 0.22 | 0.40 | 0.57 | 0.40 |
| Polymicrobial\|Diabetes | 0.29 | 0.73 | 1.14 | 0.72 |
| Polymicrobial\|Is.betalactam.inhibitor | 0.34 | 0.72 | 1.09 | 0.71 |
| CLI\|(Intercept) | -1.02 | -0.85 | -0.68 | -0.85 |
| CLI\|Is.other | 0.68 | 1.18 | 1.70 | 1.18 |
| Fusid\|(Intercept) | -3.99 | -3.47 | -3.03 | -3.45 |
| Fusid\|Diabetes | 0.44 | 1.21 | 1.96 | 1.23 |
| MUP\|(Intercept) | -5.07 | -4.26 | -3.60 | -4.22 |
| MUP\|Polymicrobial | 0.57 | 1.27 | 1.99 | 1.25 |
| MUP\|Fusid | 0.98 | 1.97 | 2.87 | 1.97 |
| MUP\|OXA | 0.64 | 1.33 | 2.00 | 1.31 |
| OXA\|(Intercept) | -1.51 | -1.29 | -1.08 | -1.28 |
| OXA\|Days.Hospitalized | 0.20 | 0.37 | 0.52 | 0.36 |

| | | | | |
|---|---|---|---|---|
| OXA\|CLI | 0.44 | 0.80 | 1.13 | 0.79 |
| | OR | | | |
| | 2.50% | median | 97.50% | mode |
| Dementia\|Age | 9.75 | 95.00 | 1,977.97 | 71.47 |
| Diabetes\|Age | 1.31 | 1.63 | 2.03 | 1.63 |
| CRF\|Age | 2.31 | 4.56 | 9.55 | 4.39 |
| CRF\|Diabetes | 8.77 | 20.75 | 52.60 | 19.40 |
| Is.cephalosporin\|Age | 1.16 | 1.36 | 1.58 | 1.35 |
| Is.betalactam\|Age | 1.25 | 1.52 | 1.83 | 1.51 |
| Is.betalactam.inhibitor\|Age | 1.26 | 1.53 | 1.85 | 1.52 |
| Is.aminoglycoside\|Age | 1.31 | 2.10 | 3.44 | 2.06 |
| Nosocomial\|Is.cephalosporin | 5.81 | 9.05 | 14.09 | 8.95 |
| Nosocomial\|Is.betalactam | 3.49 | 5.44 | 8.34 | 5.36 |
| Nosocomial\|Days.Hospitalized | 0.34 | 0.44 | 0.55 | 0.44 |
| Polymicrobial\|Age | 1.25 | 1.50 | 1.76 | 1.49 |
| Polymicrobial\|Diabetes | 1.34 | 2.08 | 3.11 | 2.06 |
| Polymicrobial\|Is.betalactam.inhibitor | 1.41 | 2.06 | 2.97 | 2.04 |
| CLI\|Is.other | 1.97 | 3.25 | 5.48 | 3.26 |
| Fusid\|Diabetes | 1.55 | 3.36 | 7.08 | 3.41 |
| MUP\|Polymicrobial | 1.77 | 3.55 | 7.33 | 3.48 |
| MUP\|Fusid | 2.67 | 7.15 | 17.73 | 7.18 |
| MUP\|OXA | 1.89 | 3.79 | 7.37 | 3.70 |
| OXA\|Days.Hospitalized | 1.22 | 1.44 | 1.68 | 1.44 |
| OXA\|CLI | 1.55 | 2.23 | 3.11 | 2.21 |

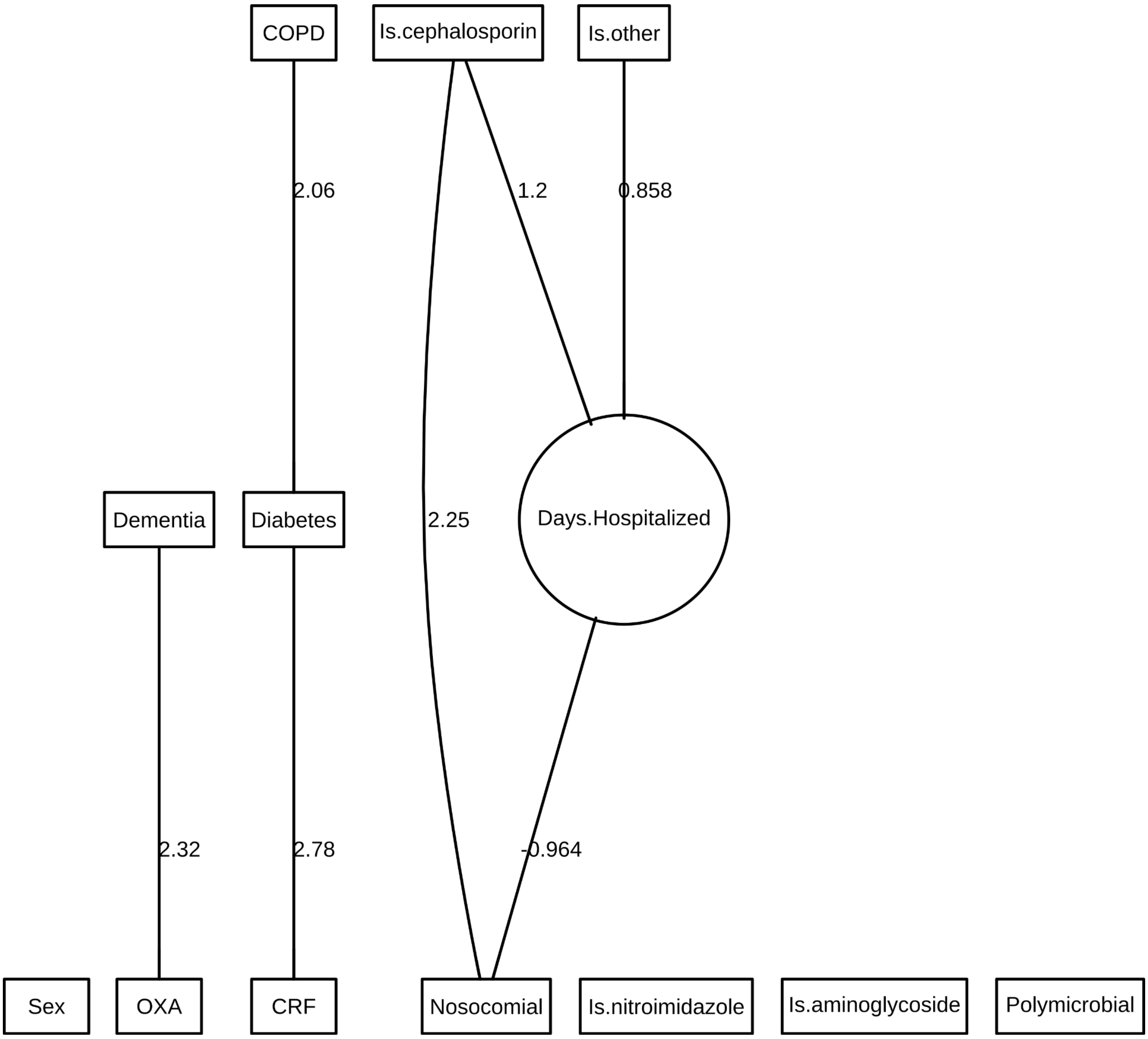

Figure S14: Final DAG for *S aureus* in aerobic blood.

Table S16: *S aureus* in aerobic blood parameter estimates and their 95% credible intervals. None of the non-intercept credible intervals contains zero.

| | Posterior statistics | | | |
|---|---|---|---|---|
| myvec | 2.50% | median | 97.50% | mode |
| Sex\|(Intercept) | 0.09 | 0.43 | 0.76 | 0.42 |
| Dementia\|(Intercept) | -3.99 | -3.09 | -2.41 | -3.04 |
| Diabetes\|(Intercept) | -2.44 | -1.90 | -1.44 | -1.88 |
| Diabetes\|COPD | 0.73 | 2.08 | 3.43 | 2.06 |
| COPD\|(Intercept) | -3.31 | -2.60 | -2.02 | -2.56 |
| CRF\|(Intercept) | -4.08 | -3.06 | -2.33 | -3.02 |
| CRF\|Diabetes | 1.65 | 2.85 | 4.04 | 2.78 |
| Is.cephalosporin\|(Intercept) | -1.06 | -0.70 | -0.38 | -0.70 |

| | | | | |
|---|---|---|---|---|
| Is.nitroimidazole\|(Intercept) | -3.06 | -2.40 | -1.86 | -2.38 |
| Is.aminoglycoside\|(Intercept) | -3.44 | -2.70 | -2.10 | -2.67 |
| Is.other\|(Intercept) | -3.06 | -2.40 | -1.86 | -2.38 |
| Days.Hospitalized\|(Intercept) | -0.62 | -0.47 | -0.33 | -0.47 |
| Days.Hospitalized\|Is.cephalosporin | 0.93 | 1.19 | 1.46 | 1.20 |
| Days.Hospitalized\|Is.other | 0.39 | 0.85 | 1.31 | 0.86 |
| Days.Hospitalized\|precision | 1.46 | 1.88 | 2.32 | 1.89 |
| Nosocomial\|(Intercept) | -1.83 | -1.27 | -0.76 | -1.23 |
| Nosocomial\|Is.cephalosporin | 1.23 | 2.32 | 3.56 | 2.25 |
| Nosocomial\|Days.Hospitalized | -1.66 | -1.00 | -0.49 | -0.96 |
| Polymicrobial\|(Intercept) | -3.31 | -2.60 | -2.02 | -2.56 |
| OXA\|(Intercept) | -1.86 | -1.41 | -1.02 | -1.40 |
| OXA\|Dementia | 0.73 | 2.45 | 4.55 | 2.32 |
| | OR | | | |
| | 2.50% | median | 97.50% | mode |
| Diabetes\|COPD | 2.08 | 8.02 | 30.96 | 7.82 |
| CRF\|Diabetes | 5.22 | 17.20 | 56.69 | 16.08 |
| Nosocomial\|Is.cephalosporin | 3.41 | 10.20 | 35.24 | 9.47 |
| Nosocomial\|Days.Hospitalized | 0.19 | 0.37 | 0.61 | 0.38 |
| OXA\|Dementia | 2.07 | 11.60 | 94.55 | 10.15 |

Table S17: Median difference and confidence intervals between parameter estimates (ln(OR)) obtained from models of bacterial species cultured from different sources.

| | | Confidence interval of difference | | | |
|---|---|---|---|---|---|
| | | 2.50% | 50% | 97.50% | |
| ***E coli*** | | | | | |
| CAZ,GEN | | | | | |
| | Urine - Wound | -0.16 | 0.06 | 0.27 | NS |
| CAZ,OFX | | | | | |
| | Aerobic - Urine | -0.25 | 0.09 | 0.44 | NS |
| | Aerobic - Wound | 0.87 | 1.00 | 1.15 | |
| | Urine - Wound | 0.71 | 0.92 | 1.12 | |
| GEN,OFX | | | | | |

| | | | | | |
|---|---|---|---|---|---|
| | Aerobic - Urine | 0.86 | 1.33 | 1.87 | |
| | Aerobic - Wound | 0.70 | 0.97 | 1.31 | |
| | Urine - Wound | -0.56 | -0.36 | -0.15 | |
| ***K pneumoniae*** | | | | | |
| GEN,OFX | | | | | |
| | Aerobic - Urine | 0.95 | 2.05 | 3.66 | |
| | Aerobic - Wound | 0.58 | 1.24 | 2.34 | |
| | Urine - Wound | -1.33 | -0.81 | -0.37 | |
| OFX,TZP | | | | | |
| | Urine - Wound | -1.46 | -0.78 | -0.27 | |
| ***P aeruginosa*** | | | | | |
| CAZ,GEN | | | | | |
| | Urine - Sputum | 0.42 | 0.57 | 0.76 | |
| | Wound - Sputum | 1.48 | 1.50 | 1.68 | |
| | Urine - Wound | -1.25 | -0.93 | -0.74 | |
| ***P Mirabilis*** | | | | | |
| Urine - Wound | | | | | |
| | AMC,CXM | 0.63 | 0.70 | 0.84 | |
| | CXM,GEN | 0.83 | 1.04 | 1.37 | |
| | CXM,OFX | -0.26 | -0.14 | -0.01 | NS* |
| | GEN,OFX | 0.64 | 0.78 | 1.06 | |

Note: AMC, amoxicillin/clavulanate; CAZ, ceftazidime; CXM, cefuroxime; GEN, gentamicin; OFX, ofloxacin; TZP, piperacillin/tazobactam. NS denotes no significant difference in parameter estimate between bacterial sources. *Effectively not significant.
